# Integrating patient experiences of healthcare: a critical realist thematic analysis of peri-operative experiences

**DOI:** 10.1101/2024.12.12.24318892

**Authors:** Neil Meigh, Liam O’Callaghan, Adrian Goldsworthy, Courtney Allan, Alison Stokes, Sharon Mickan

## Abstract

**Background:** The perioperative experience is central to how patients perceive and navigate their care. While existing research has explored broad themes, the nuanced interplay of emotional, psychological, and clinical dimensions remains underexamined. Building on a recent scoping review, this qualitative study sought to explore perioperative patient experiences in greater depth, investigating both pre-established themes and emergent insights.

**Methods:** Patients were invited to share their perioperative journeys through semi-structured interviews, offering detailed accounts of their lived experiences. Using a Critical Realist approach, the study examined both observable factors and underlying mechanisms shaping these experiences. By integrating pre-established themes with participants’ narratives, the study constructed a comprehensive framework reflecting the complexity of patient-reported experiences.

**Analysis:** Four central themes were identified: (1) psychological resilience as a central pillar for patient experience, (2) empowerment and information flow as drivers of patient experience, (3) personalised care as the foundation for patient-centred experience, and (4) integrated care and pain management as central to patient experience. Additionally, social influences and financial barriers emerged as factors shaping the broader context of these experiences, though they were less interconnected with the core themes.

**Discussion:** This study underscores the multifaceted nature of perioperative patient experiences, where emotional resilience, empowerment, and personalised care are deeply intertwined. These findings highlight the need for health systems and healthcare professionals to address emotional and contextual factors alongside clinical dimensions to improve the overall patient experience.

**Conclusion:** By prioritising the exploration of patient-reported perioperative experiences, this study provides critical insights into the emotional, psychological, and clinical elements that shape care. The integration of these dimensions offers practical guidance for developing patient-centred approaches that align with individuals’ needs and expectations.

## Introduction

The perioperative period is a critical phase in a patient’s healthcare journey, profoundly shaping their perceptions of care and influencing both immediate satisfaction and long-term outcomes. Research consistently identifies factors such as realistic expectations, accurate information, and psychological support as central to positive perioperative experiences (Accardi-Ravid et al., 2020; Anakwe et al., 2011). However, significant gaps persist between the care patients expect and what they receive, particularly regarding communication and emotional support (Auquier et al., 2005; Bovonratwet et al., 2021). These disconnects often leave patients feeling unprepared and unsupported, highlighting the importance of exploring the relational dimensions of care to complement its clinical aspects.

Beyond functional concerns, patients frequently describe the emotional and psychological dimensions of their perioperative experiences as deeply impactful. Feelings of unpreparedness for postoperative realities, including pain management and uncertain recovery timelines, are commonly reported (Van Der Horst et al., 2019). Unmet expectations can contribute to psychological distress, further complicating the recovery process and disengaging patients from their own care (Eberhart et al., 2020). As Doyle et al. (2013) demonstrated, failing to address emotional needs undermines not only satisfaction, but also clinical outcomes and safety. Preoperative anxiety, a well-documented phenomenon, exacerbates these challenges by heightening psychological distress and increasing the risk of adverse outcomes (Jovanovic et al., 2022). These accounts underscore the profoundly human nature of perioperative experiences and the necessity of integrating emotional and psychological support into clinical care.

Pain management is another pivotal element of perioperative care. While multimodal strategies have shown promise in enhancing recovery and satisfaction, many patients express dissatisfaction with their pain relief, citing unmet expectations and the absence of personalised approaches (Gobbo et al., 2020; Rejeh & Vaismoradi, 2010). These findings highlight the importance of aligning care with patients’ individual pain tolerances, cultural expectations, and broader values to foster trust and optimise the patient experience.

Non-clinical factors such as care coordination, privacy, and family involvement also shape perioperative experiences. Care coordination, encompassing the integration of services and communication among providers, has been shown to enhance patient satisfaction and reduce adverse outcomes (Manary et al., 2013; Price et al., 2014). However, much of the existing research addresses these factors in isolation, leaving critical gaps in understanding their interplay. To address this, a more holistic exploration of the perioperative experience is required, one that captures the intersection of clinical, emotional, and social dimensions.

A scoping review by Mickan et al. (2024) synthesised existing evidence on perioperative experiences, identifying four key themes: realistic expectations, accurate information, consistent communication, and individualised care. Additionally, the review highlighted four clinician focus areas that influence the patient’s perioperative experience: careful monitoring around pain management, psychological recovery, coordination of care, and return to function. While these findings provide a foundational framework for understanding perioperative care, the review underscored the need for further exploration of how these factors intersect and adapt within contemporary healthcare settings.

Building on this previous work, the present qualitative study seeks to deepen understanding of perioperative patient experiences. By amplifying patients’ voices through semi-structured interviews, this research investigates how emotional, psychological, and clinical dimensions converge to shape the perioperative journey. Guided by a Critical Realist approach, this study aims to reveal the underlying mechanisms driving these experiences, providing a framework for developing holistic, patient-centred care that foregrounds the lived experiences of those undergoing surgery.

## Methodology

### Study Design

This qualitative study utilised semi-structured interviews to explore perioperative patient experiences through a Critical Realist lens. The study aimed to capture both pre-established factors (2024) and emergent themes from participants’ narratives, offering a comprehensive understanding of these experiences. The interview guide (Supplementary File 1) was informed by Mickan et al.’s framework of patient experience factors and clinician points of influence, ensuring alignment with existing themes while encouraging participants to share novel insights. Key dimensions of care—including patient expectations, communication, and recovery—were explored through a relational approach that prioritised participant-led narratives and the co-construction of meaning.

Guided by the Critical Realist paradigm, the study examined both observable experiences and underlying mechanisms influencing perioperative care. Adherence to the BQQRG reporting guidelines for qualitative research (Braun & Clarke, 2024) is detailed in Supplementary File 2. Ethical approval was granted by the Bond University Human Research Ethics Committee [SM03248].

### Participants

Participants were recruited through Beyond The Clinic (BTC), a partner organisation collaborating with Bond University. Inclusion criteria required participants to have undergone elective surgery in Australia within the past two years. Demographic information and eligibility were assessed using a Qualtrics questionnaire (Supplementary File 3). Eligible participants were invited to one-on-one Zoom interviews, conducted by the lead researcher (NM) between 17 July and 29 September 2023.

Recruitment continued until the diversity and richness of participant narratives sufficiently addressed the study aims, ensuring exploration of both pre-established and emergent themes. This approach aligned with qualitative principles of information power, prioritising data depth and quality over sample size.

NM conducted the interviews with reflexive awareness of his clinical background, adopting a respectful and empathetic stance to mitigate potential influence. This reflexive approach balanced professional expertise with genuine curiosity, fostering an environment where participants felt empowered to share their perioperative experiences fully.

Interviews lasted 40 to 75 minutes, concluding when participants indicated they had fully expressed their perspectives. This flexible and participant-driven approach ensured the collection of rich, personal, and contextually grounded accounts of perioperative experiences.

### Data Collection and Transcription

Semi-structured interviews provided participants with the flexibility to reflect on and articulate their perioperative experiences in a conversational, participant-led manner. This qualitative approach prioritised the co-construction of meaning, allowing participants to shape the dialogue and share experiences in ways that felt authentic and personally significant. The interview guide offered a flexible framework to ensure alignment with pre-established themes while accommodating the emergence of novel insights.

All interviews were audio-recorded with participants’ informed consent, and pseudonyms were assigned to ensure confidentiality. Transcribed interviews followed an intelligent verbatim approach, omitting filler words and false starts while preserving the essential meaning, nuance, and emotive content of participants’ accounts (Eftekhari, 2024). This method aligned with qualitative principles emphasising the depth and contextual richness of participant narratives over textual precision (Braun & Clarke, 2024).

To ensure data accuracy and trustworthiness, transcribed interviews were independently reviewed against the audio recordings by two investigators (NM, LO). This process safeguarded fidelity to participants’ narratives, reinforced reflexive engagement, and incorporated multiple perspectives to enhance the credibility of the dataset.

### Thematic Analysis Using the Four C’s Model

Thematic analysis employed Christodoulou’s Four C’s model (Christodoulou, 2023), a structured framework encompassing four stages: Coding, Clustering, Connecting, and Constructing (see Table 1). This model was selected to align with the study’s goal of exploring both pre-established factors and emergent insights, facilitating a comprehensive examination of how these dimensions interact within perioperative experiences. The Four C’s approach prioritised detailed exploration of the data, ensuring a nuanced understanding of the relationships and patterns shaping participants’ accounts.

**Table 1:** The four C’s model of thematic analysis.

| Stage | Description | Key Processes |
| --- | --- | --- |
| Stage 1:<br>**Coding** | Text segments were systematically coded to capture pre-established factors (e.g., realistic expectations, communication) and emergent insights from participant narratives. | Pre-established codes were applied, and new codes were reflexively developed to reflect novel dimensions of patient experiences, ensuring contextual depth. |
| Stage 2:<br>**Clustering** | Codes were grouped into categories by assessing alignment and divergence between pre-established and emergent codes, merging similar concepts while retaining distinct categories. | Reflexive team discussions ensured that clustering retained the complexity of participant experiences. Overlap and alignment were evaluated iteratively. |
| Stage 3:<br>**Connecting** | Social network analysis revealed relationships between categories, with '1' indicating meaningful connections and '0' denoting no influence. This visualisation identified interactions shaping experiences. | A one-mode network matrix visualised category relationships, enabling the identification of enabling, conditional, and reciprocal influences across emotional, clinical, and psychological dimensions. |
| Stage 4:<br>**Constructing** | Overarching themes were developed by synthesising the strongest category connections, identifying central and autonomous influences, and hypothesising underlying mechanisms. | Hierarchical structures reflected category connectivity, providing a coherent framework for understanding the multidimensional influences shaping perioperative experiences. |

A key feature of the Four C’s model is its use of ‘categories’ rather than ‘themes’ during the clustering phase, reflecting a granular focus that aligns with the study’s Critical Realist methodology. This distinction enabled the analysis to capture nuanced connections between specific aspects of the perioperative experience while complementing the broader thematic analysis conducted in the scoping review. By iteratively engaging with the data, the analysis identified relational patterns, enabling deeper interpretation of participants’ narratives.

The model integrates retroductive reasoning to explore underlying causal mechanisms shaping participant experiences. This approach allowed the study to move beyond surface-level patterns, linking observed phenomena to systemic and relational drivers. By systematically navigating the four analytical stages, the methodology maintained coherence and flexibility, capturing the depth and complexity of perioperative experiences. This iterative process provided a multidimensional framework for understanding the interplay between emotional, psychological, and clinical factors shaping patients’ journeys.

### Critical Realist Approach to Thematic Analysis

This study adopted a Critical Realist (CR) approach to thematic analysis, complementing descriptive and interpretive frameworks by delving into the underlying mechanisms shaping perioperative experiences. The CR paradigm was selected to explore both relational dimensions (e.g., emotional and psychological aspects) and functional dimensions (e.g., clinical and procedural factors), offering a multidimensional lens through which to interpret participants’ narratives.

The CR approach facilitated an investigation not only of what patients described but also of why and how these experiences occurred, aligning with its emphasis on uncovering complexity and interconnectedness (Fletcher, 2017). Observable phenomena—such as communication, pain management, and care coordination—were analysed in relation to underlying drivers, including psychological resilience and social influences. This dual focus enabled the study to situate participants’ accounts within broader systemic and contextual dynamics, deepening understanding of their perioperative journeys.

A core element of the CR methodology is retroductive reasoning, which seeks to hypothesise and explain causal mechanisms that underpin observed patterns in the data. By interrogating the structures and conditions shaping perioperative experiences, the study identified both pre-established factors and emergent dimensions of care. This approach integrated the insights from thematic analysis into a cohesive interpretive framework, connecting individual experiences to broader relational and systemic processes. Through this lens, the study offers actionable insights for improving patient-centred perioperative care.

### Integration of Pre-established and Emergent Factors

This study synthesised pre-established patient experience factors and clinician points of influence (Mickan et al., 2024) with emergent insights derived from patient interviews, creating a unified thematic framework. Emergent categories that could not be aligned with pre-existing factors were systematically organised into new categories, enabling a comprehensive comparison that refined both anticipated and novel dimensions of perioperative experiences.

Three investigators (NM, AG, LO) engaged in iterative and reflexive discussions to review and refine the clustering process. Overlapping categories were merged to enhance coherence, while distinct categories were preserved to honour the complexity and nuance of participants’ narratives. This process resulted in a final set of ten unified categories that integrate both pre-established and emergent insights, capturing the multifaceted nature of perioperative patient experiences.

### Data Saturation and Trustworthiness

This study adopted Braun and Clarke’s (2021) approach to data saturation, treating meaning generation as an iterative and evolving process rather than aiming for a definitive endpoint where no new themes emerge. Interviews concluded when participants indicated they had fully expressed their experiences, ensuring a participant-led approach that prioritised narrative depth and authenticity.

To ensure rigour, reflexivity was embedded throughout the research process, with the team critically reflecting on potential biases and assumptions. Investigator triangulation involved independent data analysis by three researchers (NM, AG, LO), enriching the interpretive process through diverse perspectives. Peer debriefing sessions facilitated collaborative discussions, resolving discrepancies and enhancing analytical transparency. Thick descriptions of codes and categories captured the contextual and emotional complexity of perioperative experiences, ensuring that the findings authentically reflected participants’ voices and lived realities.

## Analysis

This study engaged with 19 participants who had recently undergone a diverse range of elective surgical procedures (Table 2). Participants described experiences across various life stages and healthcare needs, with surgeries ranging from planned Caesarean sections and cancer-related procedures to orthopaedic interventions and dental extractions. Their hospital stays varied from single-day procedures to extended admissions of up to 14 days, reflecting diverse perioperative experiences.

**Table 2:** Participant characteristics.

| ID | Age group | Sex | Type of Surgery | Time Since Surgery | Hospital Setting | Length of Stay | Preoperative Anxiety Levels | Pain Management Satisfaction | Family Involvement in Care |
| --- | --- | --- | --- | --- | --- | --- | --- | --- | --- |
| 1 | 30–39 | F | Planned Caesarean (C-Section) | < 3 months | Private | 5 days | Low | Satisfied | Yes |
| 2 | 20–29 | F | Full Dental Extraction | < 3 months | Private | Day surgery | High | Dissatisfied | Yes |
| 3 | 70–79 | M | Blood Clot Removal | < 3 months | Public | 3 days | High | Satisfied | No |
| 4 | 60–69 | F | Ganglion Excision (Ankle) | < 3 months | Private | Day surgery | Moderate | Dissatisfied | Yes |
| 5 | 70–79 | M | Prostate Cancer Removal | < 6 months | Private | 2 days | High | Satisfied | Yes |
| 6 | 50–59 | M | Eye Surgery (Trauma-related) | < 12 months | Public | 7 days | High | Satisfied | No |
| 7 | 70–79 | F | Rectorrhaphy | < 6 months | Private | 5 days | Low | Satisfied | No |
| 8 | 30–39 | F | Achilles Z-Lengthening & Bilateral Gastrocnemius Reduction | < 6 months | Public | 3 days | Moderate | Satisfied | Yes |
| 9 | 60–69 | M | TURP (Prostate Surgery) & Inguinal Hernia Repair | < 18 months | Public | 1 day (hernia); 2 days (TURP) | Moderate | Satisfied | Yes |
| 10 | 70–79 | F | Screw Removal (Bunion Surgery) | < 12 months | Private | Day surgery | High | Dissatisfied | No |
| 11 | 60–69 | F | Rotator Cuff Repair | < 6 months | Private | 1 day | Moderate | Satisfied | Yes |
| 12 | 30–39 | F | Planned Caesarean (C-Section) | < 6 months | Private | 5 days | Low | Satisfied | Yes |
| 13 | 70–79 | M | Bowel Surgery (Twisted Bowel) | < 2 months | Public | 5 days | High | Satisfied | Yes |
| 14 | 60–69 | F | Radical Surgery for Vulval Cancer with Colostomy | < 24 months | Public | 14 days | High | Satisfied | Yes |
| 15 | 40–49 | F | Mastectomy with DIEP Reconstruction & Lymph Node Removal | < 12 months | Private | 5 days | Moderate | Satisfied | Yes |
| 16 | 50–59 | M | Varicose Vein Surgery | < 12 months | Public | Day surgery | Low | Satisfied | No |
| 17 | 60–69 | M | Oral Cancer Removal with Reconstruction | < 6 months | Public | 7 days | High | Satisfied | Yes |
| 18 | 30–39 | F | Hemi-Thyroidectomy | < 6 months | Private | 1 day | Low | Satisfied | Yes |
| 19 | 50–59 | F | Hip Replacement | < 6 months | Public | 7 days | High | Dissatisfied | Yes |

Participants shared accounts of their emotional and psychological experiences during the perioperative period. Preoperative anxiety ranged from mild to significant, shaping their preparedness and expectations for surgery. Their reflections on pain management revealed a spectrum of satisfaction, with some participants recounting frustration or unmet expectations and others highlighting moments of relief or support. Across many narratives, the involvement of family members emerged as integral, shaping patients’ emotional resilience and perceptions of support throughout their perioperative journey.

### Overview of Integrated Categories

The analysis identified ten categories that reflect key dimensions of the patient experience during the perioperative period. These categories, presented in Table 3, draw from both pre-established factors and emergent insights derived from participants’ narratives. Together, they offer a holistic view of the multifaceted experiences reported by patients, capturing their emotional, psychological, and relational dimensions alongside clinical interactions.

**Table 3:**
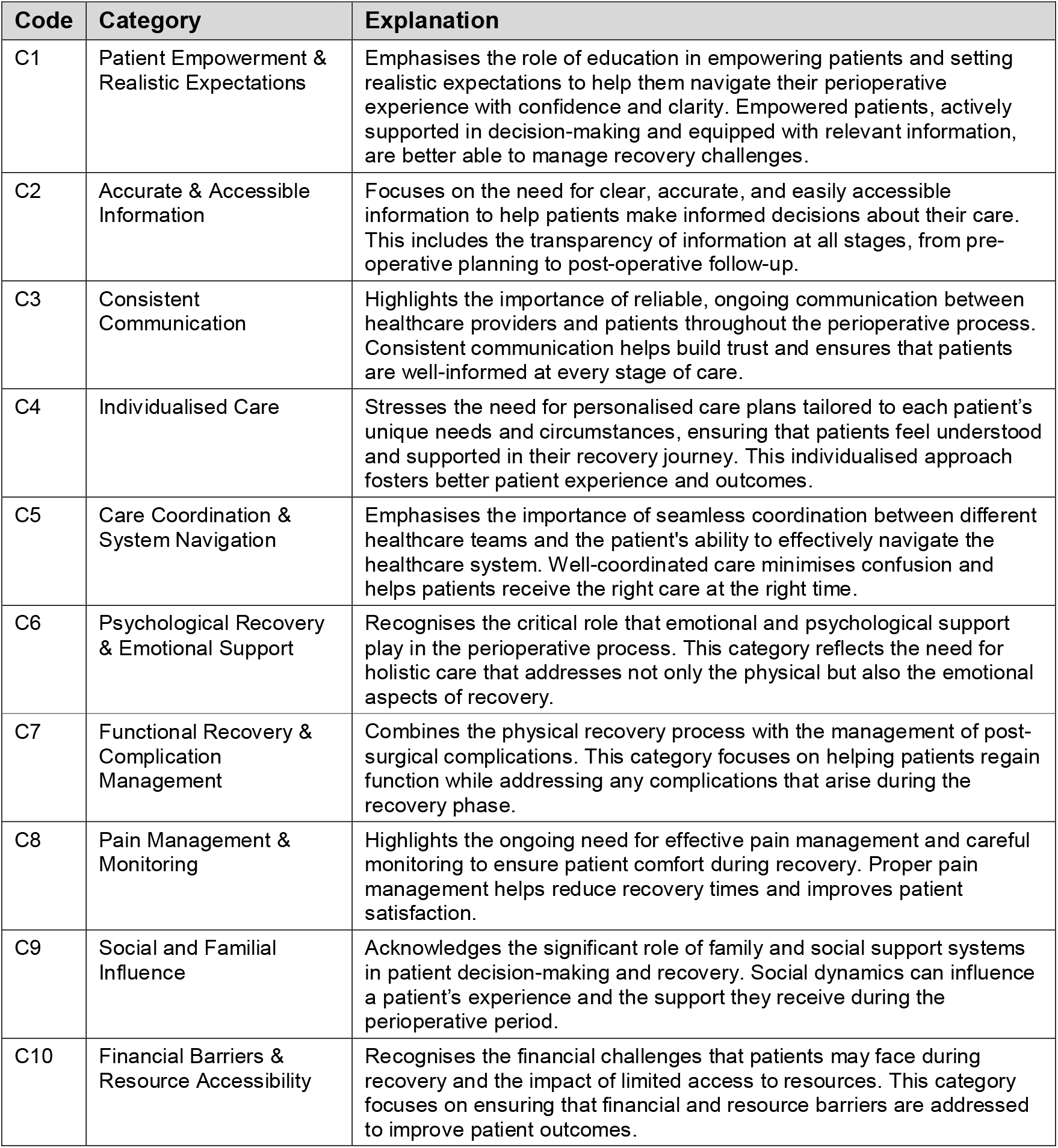
Summary of unified categories impacting perioperative patient experiences.

| Code | Category | Explanation |
| --- | --- | --- |
| C1 | Patient Empowerment & Realistic Expectations | Emphasises the role of education in empowering patients and setting realistic expectations to help them navigate their perioperative experience with confidence and clarity. Empowered patients, actively supported in decision-making and equipped with relevant information, are better able to manage recovery challenges. |
| C2 | Accurate & Accessible Information | Focuses on the need for clear, accurate, and easily accessible information to help patients make informed decisions about their care. This includes the transparency of information at all stages, from pre-operative planning to post-operative follow-up. |
| C3 | Consistent Communication | Highlights the importance of reliable, ongoing communication between healthcare providers and patients throughout the perioperative process. Consistent communication helps build trust and ensures that patients are well-informed at every stage of care. |
| C4 | Individualised Care | Stresses the need for personalised care plans tailored to each patient's unique needs and circumstances, ensuring that patients feel understood and supported in their recovery journey. This individualised approach fosters better patient experience and outcomes. |
| C5 | Care Coordination & System Navigation | Emphasises the importance of seamless coordination between different healthcare teams and the patient's ability to effectively navigate the healthcare system. Well-coordinated care minimises confusion and helps patients receive the right care at the right time. |
| C6 | Psychological Recovery & Emotional Support | Recognises the critical role that emotional and psychological support play in the perioperative process. This category reflects the need for holistic care that addresses not only the physical but also the emotional aspects of recovery. |
| C7 | Functional Recovery & Complication Management | Combines the physical recovery process with the management of post-surgical complications. This category focuses on helping patients regain function while addressing any complications that arise during the recovery phase. |
| C8 | Pain Management & Monitoring | Highlights the ongoing need for effective pain management and careful monitoring to ensure patient comfort during recovery. Proper pain management helps reduce recovery times and improves patient satisfaction. |
| C9 | Social and Familial Influence | Acknowledges the significant role of family and social support systems in patient decision-making and recovery. Social dynamics can influence a patient's experience and the support they receive during the perioperative period. |
| C10 | Financial Barriers & Resource Accessibility | Recognises the financial challenges that patients may face during recovery and the impact of limited access to resources. This category focuses on ensuring that financial and resource barriers are addressed to improve patient outcomes. |

The relationships between these categories are illustrated in a one-mode network (Figure 1), visualising how interconnected aspects of the perioperative experience shape patients’ perceptions and narratives. This network provides the foundation for broader themes (Figure 2), which reflect causal mechanisms and the complexities underpinning patients’ experiences.

**Figure 1:**
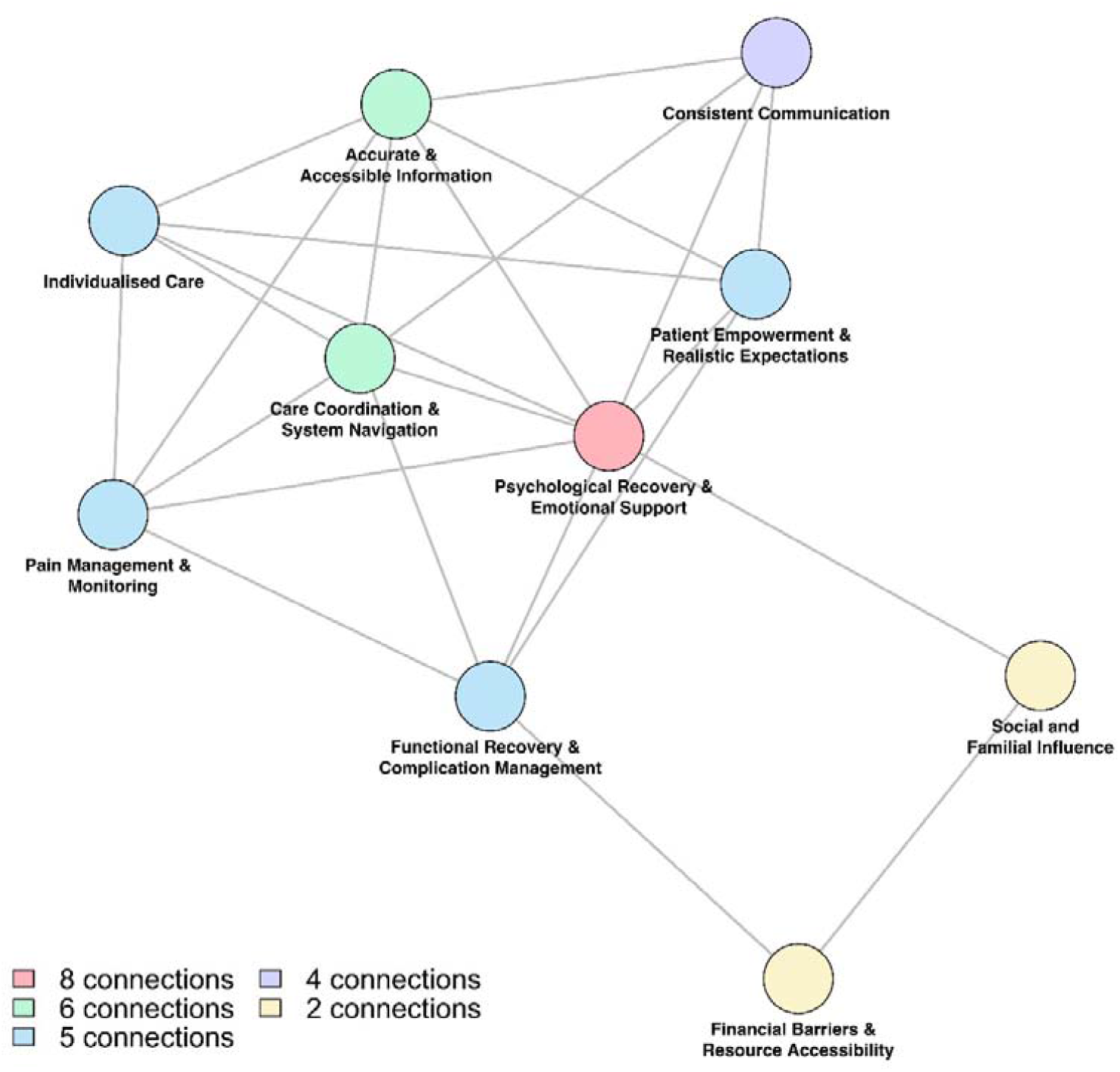
Relational map depicting category connections.

**Figure 2:**
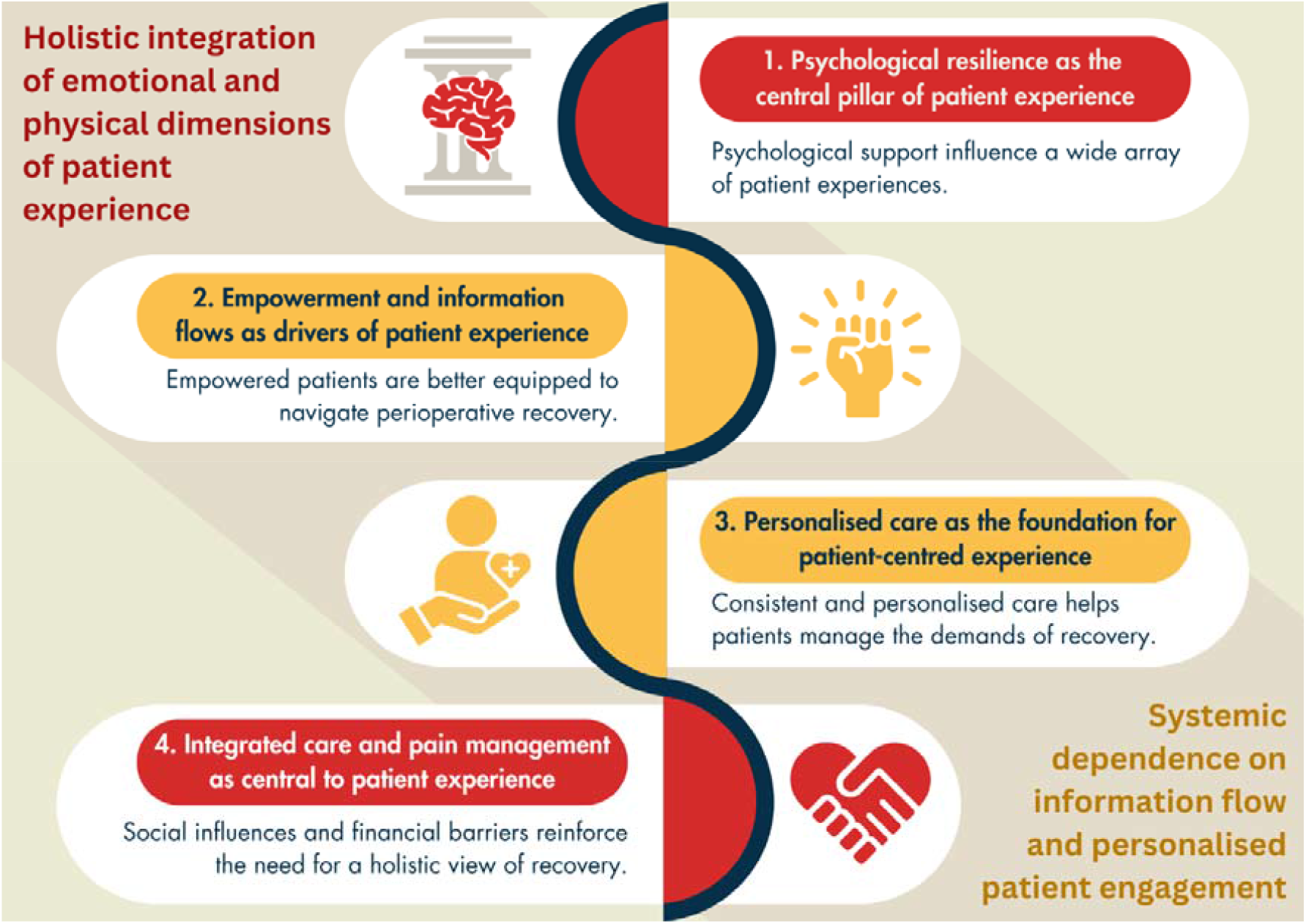
Integrated themes and causal mechanisms in perioperative patient experience.

### Stage 1 – Initial / In vivo Coding

The analysis began with an inductive coding process, independently examining text fragments from the 19 transcribed interviews. This participant-led approach identified 487 unique codes, capturing the breadth and nuance of perioperative experiences without reference to pre-existing frameworks. After this initial coding, the codes were retrospectively compared and mapped to the eight pre-established categories identified in Mickan et al.’s (2024) scoping review:

- Accurate Information: 62 codes
- Careful Monitoring around Pain Management: 49 codes
- Consistent Communication: 74 codes
- Coordination of Care: 57 codes
- Individualised Care: 58 codes
- Psychological Recovery: 96 codes
- Realistic Expectations: 45 codes
- Return to Function: 46 codes

This mapping process identified an additional 180 codes that diverged from the pre-established categories. These emergent codes represented critical aspects of patient experiences not fully addressed by the original framework, enriching the analysis and highlighting novel dimensions of perioperative care.

### Stage 2 – Clustering

The clustering stage organised codes into both pre-established and emergent categories, providing a structured foundation for understanding perioperative patient experiences. Seven new categories, developed from patient-reported narratives, were evaluated alongside pre-established categories to identify overlaps, refine definitions, and minimise duplication. This iterative process integrated emergent and pre-established insights into a final set of ten unified categories, presented in Table 3.

The clustering of codes into pre-established and emergent categories are presented in Supplementary File 4 and File 5, respectively, while representative text segments illustrating the emotional and personal dimensions of these experiences are provided in Supplementary File 6. The mapping and integration of pre-established and emergent categories, shown in Supplementary File 7, highlight the development of the final unified categories. Two entirely new categories emerged from the analysis: *Social and Familial Influence* and *Financial Barriers & Resource Accessibility*, reflecting critical external factors shaping perioperative experiences.

This final set of categories reflects a comprehensive framework encompassing clinical, emotional, psychological, and social dimensions. By integrating patient-reported experiences and pre-established insights, the unified categories offer a robust foundation for exploring the relational and causal mechanisms in subsequent stages of analysis.

### Stage 3 – Connecting

A one-mode network (supplementary file 8) was constructed to examine the relationships among the ten unified categories identified in the analysis. Figure 1 visualises these connections, providing a dynamic perspective on the interconnected dimensions of perioperative patient experiences. This relational map revealed key categories central to shaping patient experiences, as determined by the number of connections.

This analysis underscores the interdependent nature of these categories, with psychological recovery, patient empowerment, and accessible information functioning as central nodes within the network.

Participants who received robust psychological support and timely, accurate information reported feeling more engaged and better prepared to navigate recovery challenges. Conversely, contextual dimensions such as Social and Familial Influence (C9) and Financial Barriers & Resource Accessibility (C10) emerged as influential factors shaping decision-making and overall experiences, extending the scope of perioperative care beyond clinical interventions to encompass broader social and economic considerations.

Supplementary File 9 provides a detailed analysis of the relational dynamics within the network, clarifying how individual categories influence and are influenced by others. These insights reinforce the multifaceted nature of perioperative care and the need for integrated approaches addressing clinical, psychological, and socio-environmental dimensions. This network analysis not only illustrates critical interconnections but also identifies focal areas for clinical practice, enhancing patient-centred care. The subsequent sections explore the thematic insights derived from these relationships and their implications for improving perioperative experiences.

### Stage 4 – Constructing

The analysis of category connections in the one-mode network provided a hierarchical framework, guiding the construction of four core themes that capture the intricacies of patients’ perioperative experiences. The strength and number of connections between categories highlighted the dynamic interplay between various aspects of care, with certain categories emerging as pivotal in shaping how patients experienced and navigated the perioperative period.

Based on connectivity:

a. C6 (Psychological Recovery & Emotional Support) attracted the most connections (8 lines), emphasising its centrality in shaping the overall patient experience:
b. C2 (Accurate & Accessible Information) and C5 (Care Coordination & System Navigation) were highly connected (6 lines), highlighting their roles in enabling patients to actively engage with their care:
c. C1 (Patient Empowerment & Realistic Expectations), C4 (Individualised Care), C7 (Functional Recovery & Complication Management), and C8 (Pain Management & Monitoring) formed the next tier, integral in ensuring patients felt supported and equipped to face the challenges of perioperative care (five lines):
d. C3 (Consistent Communication) showed fewer but still significant connections (four lines), played a key role in ensuring patients felt informed and supported throughout their perioperative journey:
e. C9 (Social & Familial Influence) and C10 (Financial Barriers & Resource Accessibility), while less interconnected (two lines), were critical external factors influencing patients’ perioperative experiences:

The relationships between these categories informed the construction of four core themes that encapsulate the multidimensional nature of the perioperative experience. These themes reflect the integration of emotional, clinical, and systemic dimensions of care, as experienced and reported by patients. By synthesising the relational and contextual factors influencing patients’ experiences, the themes provide a holistic understanding of the perioperative journey:

#### THEME 1 (connectivity (a))

**Psychological resilience as the central pillar of patient experience** Psychological recovery and emotional support (C6) emerged as the most interconnected and foundational aspect of the perioperative experience, demonstrating extensive links with patient empowerment (C1), communication (C3), functional recovery (C7), and pain management (C8). The analysis revealed that psychological resilience played a central role in shaping patients’ experiences, influencing their capacity to navigate post-surgical challenges, engage in rehabilitation, and manage pain. This category highlights how psychological support is pivotal in fostering confidence, enabling patients to participate actively in their care, and addressing the emotional demands of recovery. Its centrality underscores its influence on the interconnected dimensions of the perioperative experience, as constructed through this analysis.

#### THEME 2 (connectivity (b))

**Empowerment and information flow as drivers of patient experience** Accurate information (C2) and care coordination and system navigation (C5) emerged as key enablers of patient empowerment, demonstrating strong connections to categories such as functional recovery (C7) and pain management (C8). The analysis revealed that when patients were empowered through clear and accessible information, they were better equipped to actively engage with their care, manage complications, and make informed decisions. The integration of accurate information and effective care coordination highlights their role in shaping patient experiences by fostering confidence, facilitating understanding, and supporting navigation through the perioperative journey

#### THEME 3 (connectivity (c))

**Personalised care as the foundation for patient-centred experience** Patient empowerment (C1) and consistent communication (C3) were closely linked with individualised care (C4) and psychological support (C6), forming the foundation of personalised perioperative care. The analysis revealed that clear communication and realistic expectations enabled patients to feel empowered and engaged, while tailored care approaches ensured that individual needs and circumstances were met. This theme highlights the importance of personalised and consistent care in fostering trust, aligning care with patient priorities, and supporting both emotional and practical aspects of their perioperative experience.

#### THEME 4 (connectivity (d) and (e))

**Integrated care and pain management as central to patient experience** Care coordination and system navigation (C5) and pain management (C8) were central to guiding patients through the complexities of their perioperative care. Analysis of their connections revealed that seamless coordination across healthcare teams and effective pain management strategies enabled patients to feel supported in managing both physical and emotional challenges. Additionally, Social and Familial Influence (C9) and Financial Barriers and Resource Accessibility (C10), while less connected to other categories, were identified as external but significant contextual factors shaping how patients experienced perioperative care. These findings underscore the need for a holistic understanding of patient experiences, which incorporates social and economic dimensions alongside clinical care.

By merging certain categories and referencing the connectivity levels (a-e), we constructed cohesive themes that encapsulate the interrelated dimensions of patient empowerment, psychological recovery, communication, and care coordination. This approach avoided over-fragmentation, ensuring that the themes are meaningful and reflective of the deeper relationships influencing patient experiences. This process aligns with the critical realist methodology, allowing for the integration of emergent patterns and highlighting the interconnected factors that shape the perioperative journey.

In the final stage of analysis, retroductive reasoning was applied to infer the underlying causal mechanisms shaping patients’ perioperative experiences. The critical realist framework enabled us to move beyond descriptive observations, using the connections established through the thematic analysis to hypothesise two primary causal mechanisms. These mechanisms provide insight into how different aspects of patient experiences—psychological resilience, pain management, information flow, and care coordination—interact to shape the way patients engage with and perceive their perioperative care.

#### Causal Mechanism 1: Holistic integration of emotional and physical dimensions of patient experience

This mechanism explains Theme 1 (psychological resilience as the central pillar of patient experience) and Theme 4 (integrated care coordination and pain management). It emphasises the inseparability of emotional and physical aspects of perioperative care, positing that psychological resilience and emotional support underpin a patient’s ability to navigate physical recovery effectively. A healthcare team that holistically addresses these interconnected dimensions provides a support structure that enhances the patient experience, fostering confidence and trust. By aligning emotional support, pain management, and care coordination, this mechanism illustrates how a patient-centred approach contributes to consistent, comprehensive care that enables patients to navigate their perioperative journeys with fewer disruptions and greater confidence.

#### Causal Mechanism 2: Systemic dependence on information flow and personalised patient engagement

This mechanism explains Theme 2 (empowerment and information flow as drivers of patient experience) and Theme 3 (personalised care as the foundation of patient-centred experience). It focuses on the pivotal role of accurate, timely, and accessible information in shaping patients’ experiences of their care. When patients are provided with clear information aligned with realistic expectations, they feel empowered to actively engage in their care and decision-making processes. Personalised care strategies, supported by consistent and transparent communication, further enhance this engagement by recognising and addressing the unique needs and expectations of each patient. This mechanism highlights a system where patient education and tailored approaches are prioritised, enabling patients to feel informed, supported, and valued throughout their perioperative experience.

These two causal mechanisms do not operate in isolation but interact to create an integrated framework for understanding patients’ experiences of perioperative care. The interplay between holistic emotional and physical dimensions of patient experience (Causal Mechanism 1) and the systemic dependence on information flow and personalised care (Causal Mechanism 2) highlights the complex yet interconnected nature of perioperative experiences. Figure 2 illustrates how the four core themes identified in this study align with and are shaped by these two interrelated causal mechanisms.

These two inferred causal mechanisms illustrate the complex interplay between psychological recovery, empowerment, care coordination, and social support, providing a cohesive framework for understanding patients’ perioperative experiences. The analysis highlights how emotional and physical dimensions of care are deeply interconnected, with each mechanism contributing to a comprehensive understanding of the factors that shape patient engagement and perceptions of their care.

This integrated approach advances theoretical insights into the multidimensional nature of patient experiences while offering practical implications for enhancing perioperative care. By addressing both functional dimensions (e.g., care coordination, pain management) and relational aspects (e.g., psychological support, patient empowerment, and consistent communication), healthcare providers can create more holistic, patient-centred pathways that align with the diverse and interwoven needs of patients during the perioperative period.

## Discussion

The findings of this study highlight the intricate and interdependent nature of perioperative experiences, where psychological resilience, patient empowerment, personalised care, and integrated care coordination emerge as central to shaping the patient experience. These elements operate within a dynamic interplay, influencing both the emotional and physical dimensions of perioperative patient experience. By exploring these interactions, the study contributes to a nuanced understanding of perioperative care, advancing prior research and offering fresh insights into the multifaceted experiences of patients.

Psychological resilience—understood as the capacity to navigate and adapt to emotional and psychological challenges—was identified as central to the perioperative experience. Consistent with prior research (Heitzmann et al., 2011; Stanton et al., 2007), our findings underscore that emotional and psychological support are foundational, not supplementary, to recovery. Participants described how psychological recovery, involving emotional healing and adaptation, intersected with other key elements of their care, including empowerment, communication, and pain management. For example, consistent with Yang et al. (2022), participants with stronger psychological resilience navigated discrepancies between preoperative expectations and postoperative realities with greater ease. Similarly, aligned with Kästner et al. (2021), participants who demonstrated psychological resilience reported being better equipped to manage pain and engage actively in rehabilitation, underscoring the interconnectedness of emotional and physical recovery.

Our findings extend this understanding by emphasising the pivotal role of psychological support within perioperative pathways. Participants who described receiving robust emotional support reported greater engagement with their care, adherence to treatment plans, and improved pain management. These observations highlight the importance of embedding psychological support within routine perioperative care to create more cohesive, patient-centred environments. In line with Kehlet and Dahl’s (2003) framework, integrating psychological care alongside physical interventions enhances the quality and consistency of recovery support.

Accurate and accessible information emerged as a key driver of patient empowerment. Participants emphasised the value of clear, timely, and relevant information in helping them navigate their care and recovery. Empowerment in this context reflects patients’ capacity to make informed decisions, engage actively in their care, and manage recovery challenges with confidence. These findings align with research demonstrating that effective communication and shared decision-making underpin positive patient experiences (Araujo et al., 2022; Pomey et al., 2015).

Care coordination, defined as the seamless integration of services and communication among healthcare providers, played an essential role in sustaining the flow of information and minimising fragmentation of care. Consistent and proactive communication emerged as a strategy to enhance patients’ sense of security and control. Participants who described experiencing coordinated care noted fewer disruptions and smoother transitions, reinforcing the need for clinicians to prioritise care integration. These observations align with Fix et al. (2018), who highlight the significance of clear communication and effective coordination in empowering patients and fostering positive recovery trajectories.

Personalised care, tailored to individual patient needs and preferences, was consistently identified as a cornerstone of positive perioperative experiences. This aligns with the person-centred frameworks described by Wolf et al. (2021), which emphasise that patient experiences are shaped by the totality of interactions with healthcare systems. Participants in our study who reported receiving personalised care noted higher levels of trust and engagement, suggesting that tailored approaches foster meaningful connections between patients and their healthcare providers. These findings also resonate with Abraham et al. (2023), who highlight the role of personalised care in building trust and enhancing recovery.

However, the study also highlights systemic constraints that can hinder personalised care delivery, particularly in resource-limited settings. Efficiency and standardisation often take precedence, limiting the adaptability of care plans. Addressing these structural barriers is critical to enabling healthcare systems to meet the diverse needs of patients while maintaining equitable care standards.

Effective care coordination and pain management emerged as dual pillars supporting successful perioperative recovery. Participants described coordinated transitions between healthcare providers as critical to ensuring continuity of care. These findings align with Kehlet and Dahl’s (2003) emphasis on the interplay between pain management and functional recovery. Adequate pain control was not only a source of comfort but also a facilitator of active participation in rehabilitation. This highlights the dual role of pain management as both a physiological necessity and a psychological enabler of recovery.

The study underscores the impact of external factors, such as social support and financial barriers, on the perioperative experience. Social networks were identified as both facilitators and barriers, reflecting the complex dynamics of family involvement. While supportive relationships provided essential encouragement and assistance, conflicting priorities within families occasionally introduced challenges to the care process. Additionally, financial barriers, such as limited access to rehabilitation resources, compounded recovery difficulties for some participants. These findings align with Reblin and Uchino’s (2008) work on the influence of financial and social factors on recovery, emphasising the need for a holistic approach to perioperative care that integrates medical and social considerations.

The two inferred causal mechanisms offer a cohesive framework for understanding the perioperative experience. Psychological resilience, central to Causal Mechanism 1, is closely linked to the empowerment and information flow described in Causal Mechanism 2. Participants who felt empowered through timely information and robust support systems were better equipped to navigate challenges and engage actively in their recovery. These mechanisms interact to create a patient-centred model of perioperative care, addressing both functional and relational dimensions. By prioritising integrated care strategies, health systems and healthcare professionals can foster environments that address the holistic needs of patients, ensuring both emotional and physical well-being throughout the perioperative journey.

This study has several limitations that warrant reflection. First, while the sample represents diverse perioperative experiences, the recruitment process may have unintentionally excluded perspectives from individuals with limited access to healthcare services or those who experienced severe complications. Additionally, the retrospective nature of participant accounts introduces the potential for recall bias. The study’s reliance on self-reported data may also reflect subjective interpretations rather than objective occurrences, though this aligns with its focus on lived experiences. Future research should aim to explore perioperative experiences in more diverse healthcare settings and examine the applicability of the proposed framework to specific surgical contexts. Despite these limitations, this study provides actionable insights for improving perioperative care by addressing both clinical and psychosocial dimensions of patient experience. By situating these insights within a Critical Realist framework, the study advances theoretical understanding while offering practical implications for enhancing patient-centred care.

## Conclusion

This study offers a nuanced framework for understanding perioperative experiences, emphasising the central roles of psychological resilience, patient empowerment, personalised care, and care coordination. By integrating these dimensions through a Critical Realist lens, we identified two primary causal mechanisms: the holistic integration of emotional and physical dimensions of the patient experience and the systemic reliance on information flow and personalised patient engagement.

These mechanisms underscore how interconnected dimensions of care shape patients’ perioperative experiences, providing a cohesive foundation for both theoretical advancement and practical intervention.

The findings emphasise that psychological and emotional support are fundamental to the perioperative experience, shaping patients’ ability to navigate challenges and engage actively in their care. Empowered patients—those provided with clear, timely information, personalised care, and robust emotional support—are better positioned to participate in recovery processes. These insights highlight the importance of aligning clinical, emotional, and systemic factors to foster cohesive perioperative pathways.

Future interventions should prioritise the integration of these elements within healthcare systems, ensuring that emotional and physical dimensions of recovery are addressed holistically. Structural flexibility is essential to enable personalised care approaches, particularly in resource-constrained settings, where standardisation often takes precedence. Tailoring care to meet individual needs will build trust, enhance engagement, and improve alignment between patients and healthcare providers.

Ultimately, this research offers a patient-centred model for perioperative care, emphasising the interplay between psychological resilience, care coordination, and empowerment through information flow. By addressing these dimensions as interdependent priorities, healthcare systems can create equitable and effective recovery pathways that respond to the lived realities of patients.

## Data Availability

All data produced in the present work are contained in the manuscript.

## Acknowledgements

The authors sincerely thank the patients who generously shared their lived experiences of hospital and peri-operative care, without whom this study would not have been possible. Thanks also to Carly Hudson, for creating ‘Figure 2’ for this manuscript.

## Author’s contributions

CRediT author statement – **Neil Meigh**: Conceptualisation, Data curation, Formal analysis, Investigation, Methodology, Project administration, Resources, Software, Validation, Writing – original draft, – review & editing. **Sharon Mickan**: Conceptualisation, Methodology, Project administration, Supervision, Validation, Writing – review & editing. **Adrian Goldsworthy**: Data Curation, Formal Analysis, Methodology, Resources, Software, Validation, Visualisation, Writing – Review & Editing. **Liam O’Callaghan**: Data curation, Formal analysis, Methodology, Software, Validation, Writing – review & editing. **Alison Stokes**: Conceptualisation, Investigation, Methodology, Project administration, Writing – review & editing. **Courtney Allan**: Conceptualisation, Investigation, Resources, Writing – review & editing.

## Availability of data and materials

All data generated or analysed during this study are included in this published article. Bond University Human Research Ethics Committee has not authorised the public release of participants’ transcribed interviews.

## Declarations

### Ethics approval and consent to participate

All research activities were conducted in accordance with relevant guidelines and regulations, in accordance with the Declaration of Helsinki. The study was approved by the Bond University Human Research Ethics Committee (BUHREC; [SM03248]). Informed consent was obtained by the lead investigator from all participants.

### Consent for publication

Not applicable.

## References

Abraham, J., Meng, A., Baumann, A., Holzer, K. J., Lenard, E., Freedland, K. E., Lenze, E. J., Avidan, M. S., & Politi, M. C. (2023). A multi– and mixed-method adaptation study of a patient-centered perioperative mental health intervention bundle. BMC Health Services Research, 23(1), 1175. 10.1186/s12913-023-10186-3

Accardi-Ravid, M. C., Eaton, L. H., Meins, A. R., Godfrey, D. S., Gordon, D. B., Lesnik, I., & Doorenbos, A. Z. (2020). A qualitative descriptive study of patient experiences of pain before and after spine surgery. Pain Medicine. 10.1093/PM/PNZ090

Anakwe, R. E., Jenkins, P. J., & Moran, M. D. (2011). Predicting dissatisfaction after total hip arthroplasty: A study of 850 patients. Journal of Arthroplasty. 10.1016/J.ARTH.2010.03.013

Araujo, C. G., de Souza e Silva, C.G., Laukkanen, J. A., Fiatarone Singh, M., Kunutsor, S. K., Myers, J., Franca, J. F., & Castro, C. L. (2022). Successful 10-second one-legged stance performance predicts survival in middle-aged and older individuals. British Journal of Sports Medicine, 56(17), 975. 10.1136/bjsports-2021-105360

Aujoulat, I., d’Hoore, W., & Deccache, A. (2007). Patient empowerment in theory and practice: Polysemy or cacophony? Patient Education and Counseling, 66(1), 13–20. 10.1016/j.pec.2006.09.008

Auquier, P., Pernoud, N., Bruder, N., Siméoni, M., Auffray, J., Colavolpe, C., François, G., Gouin, F., Manelli, J., Martin, C. D., Sapin, C., & Blache, J. (2005). Development and validation of a perioperative satisfaction questionnaire. Anesthesiology. 10.1097/00000542-200506000-00010

Bovonratwet, P., Shen, T. S., Islam, W., Sculco, P. K., Padgett, D. E., & Su, E. P. (2021). Is there an association between negative patient-experience comments and perioperative outcomes after primary total hip arthroplasty? Journal of Arthroplasty. 10.1016/J.ARTH.2021.01.023

Braun, V., & Clarke, V. (2021). To saturate or not to saturate? Questioning data saturation as a useful concept for thematic analysis and sample-size rationales. Qualitative Research in Sport, Exercise and Health, 13(2), 201–216. 10.1080/2159676X.2019.1704846

Christodoulou, M. (2023). The four C’s model of thematic analysis. A critical realist perspective. Journal of Critical Realism, 22(1), 1–20.

Clarke, V., & Braun, V. (2017). Thematic analysis. The Journal of Positive Psychology, 12(3), 297–298. 10.1080/17439760.2016.1262613

Doyle, C., Lennox, L., & Bell, D. (2013). A systematic review of evidence on the links between patient experience and clinical safety and effectiveness. BMJ Open. 10.1136/BMJOPEN-2012-001570

Eberhart, L., Aust, H., Schuster, M., Sturm, T., Gehling, M., Euteneuer, F., & Rüsch, D. (2020). Preoperative anxiety in adults—A cross-sectional study on specific fears and risk factors. BMC Psychiatry. 10.1186/S12888-020-02552-W

Eftekhari, H. (2024). Transcribing in the digital age: Qualitative research practice utilizing intelligent speech recognition technology. European Journal of Cardiovascular Nursing, 23(5), 553–560. 10.1093/eurjcn/zvae013

Fix, G. M., Lukas, C. V., Bolton, R. E., Hill, J., Mueller, N., LaVela, S. L., & Bokhour, B. G. (2018). Patient-centred care is a way of doing things: How healthcare employees conceptualize patient-centred care. 10.1111/HEX.12615

Fletcher, A. J. (2017). Applying critical realism in qualitative research: Methodology meets method. International Journal of Social Research Methodology, 20(2), 181–194. 10.1080/13645579.2016.1144401

Forsberg, A., Vikman, I., Wälivaara, B., & Engström, Å. (2015). Patients’ perceptions of quality of care during the perioperative procedure. Journal of Perianesthesia Nursing_: Official Journal of the American Society of PeriAnesthesia Nurses. 10.1016/J.JOPAN.2014.05.012

Gobbo, M., Saldaña, R., Rodríguez, M., Jiménez, J., García-Vega, M. I., Pedro, J.M.de, & Cea-Calvo, L. (2020). Patients’ experience and needs during perioperative care: A focus group study. Patient Preference and Adherence. 10.2147/PPA.S252670

Heitzmann, C. A., Merluzzi, T. V., Jean-Pierre, P., Roscoe, J. A., Kirsh, K. L., & Passik, S. D. (2011). Assessing self-efficacy for coping with cancer: Development and psychometric analysis of the brief version of the Cancer Behavior Inventory (CBI-B). Psycho-Oncology, 20(3), 302–312. 10.1002/pon.1735

Jovanovic, K., Kalezić, N., & Šipetic-Grujicic, S. (2022). Preoperative anxiety: An important, but neglected issue. Medicinska Istrazivanja. 10.5937/MEDI55-40195

Kästner, A., Ng Kuet Leong, V.S.C., Petzke, F., Budde, S., Przemeck, M., Müller, M., & Erlenwein, J. (2021). The virtue of optimistic realism—Expectation fulfillment predicts patient-rated global effectiveness of total hip arthroplasty. BMC Musculoskeletal Disorders, 22(1), 180. 10.1186/s12891-021-04040-y

Kehlet, H., & Dahl, J. B. (2003). Anaesthesia, surgery, and challenges in postoperative recovery. The Lancet, 362(9399), 1921–1928. 10.1016/S0140-6736(03)14966-5

Manary, M. P., Boulding, W., Staelin, R., & Glickman, S. W. (2013). The patient experience and health outcomes. The New England Journal of Medicine. 10.1056/NEJMP1211775

Manser, T. (2009). Teamwork and patient safety in dynamic domains of healthcare: A review of the literature. Acta Anaesthesiologica Scandinavica, 53(2), 143–151. 10.1111/j.1399-6576.2008.01717.x

Mickan, S., Fletcher, J., Burrows, R., Bateup, S., Stokes, A., & Tsung, J. (2024). Reporting patient experiences within elective perioperative care: A scoping review. International Journal for Quality in Health Care, 36(3), mzae085. 10.1093/intqhc/mzae085

Pomey, M.-P., Ghadiri, D. P., Karazivan, P., Fernandez, N., & Clavel, N. (2015). Patients as partners: A qualitative study of patients’ engagement in their health care. PLOS ONE, 10(4), e0122499. 10.1371/journal.pone.0122499

Price, R. A., Elliott, M., Zaslavsky, A., Hays, R., Lehrman, W. G., Rybowski, L., Edgman-Levitan, S., & Cleary, P. (2014). Examining the role of patient experience surveys in measuring health care quality. Medical Care Research and Review_: MCRR, 71(5), 522–554. 10.1177/1077558714541480

Reblin, M., & Uchino, B. N. (2008). Social and emotional support and its implication for health. Current Opinion in Psychiatry, 21(2), 201–205. 10.1097/yco.0b013e3282f3ad89

Rejeh, N., & Vaismoradi, M. (2010). Perspectives and experiences of elective surgery patients regarding pain management. Nursing & Health Sciences. 10.1111/J.1442-2018.2009.00488.X

Stanton, A. L., Revenson, T. A., & Tennen, H. (2007). Health psychology: Psychological adjustment to chronic disease. Annual Review of Psychology, 58(1), 565–592. 10.1146/annurev.psych.58.110405.085615

Valtorta, N. K., Kanaan, M., Gilbody, S., Ronzi, S., & Hanratty, B. (2016). Loneliness and social isolation as risk factors for coronary heart disease and stroke: Systematic review and meta-analysis of longitudinal observational studies. Heart, 102(13), 1009–1016. 10.1136/heartjnl-2015-308790

Van Der Horst, A. Y., Trompetter, H. R., Pakvis, D. F. M., Kelders, S. M., Schreurs, K. M. G., & Bohlmeijer, E. T. (2019). Between hope and fear: A qualitative study on perioperative experiences and coping of patients after lumbar fusion surgery. International Journal of Orthopaedic and Trauma Nursing, 35, 100707. 10.1016/j.ijotn.2019.07.003

Vitous, C. A., Byrnes, M. E., De Roo, A., Jafri, S. M., & Suwanabol, P. A. (2022). Exploring emotional responses after postoperative complications: A qualitative study of practicing surgeons. Annals of Surgery, 275(1), e124–e131. 10.1097/sla.0000000000004041

Wolf, J. A., Niederhauser, V., Marshburn, D., & LaVela, S. L. (2021). Reexamining “Defining Patient Experience”: The human experience in healthcare. Patient Experience Journal, 8(1), 16–29. 10.35680/2372-0247.1594

Yang, G., Shen, S., Zhang, J., & Gu, Y. (2022). Psychological resilience is related to postoperative adverse events and quality of life in patients with glioma: A retrospective cohort study. Translational Cancer Research, 11(5), 1219–1229. 10.21037/tcr-22-732

