## Supplementary File 1 for "Integrating patient experiences of healthcare: a critical realist thematic analysis of peri-operative experiences"

##### Supplementary File 1. Interview guide

| **Theme** | **Interview Questions** |
| --- | --- |
| Realistic Expectations | What were your expectations of surgery? |
|  | Was your surgeon aware of your expectations? |
|  | Do you think your expectations were realistic? |
|  | Were you given an opportunity to discuss your expectations with your healthcare team? |
| Accurate Information | Did you have any questions about your surgery? How were these answered or clarified? |
|  | What information did you receive about your surgery, and did you find it helpful? |
|  | What information were you given to take home with you after surgery? |
|  | Was the information about your surgery shared with anyone else (e.g., family, carer)? |
|  | Was the information consistent throughout your care? |
|  | Did you receive information about your surgery or recovery from other sources (e.g., patients, friends, family, the internet)? |
| Consistent Communication | Who did you communicate with about your surgery? (e.g., GP, specialist’s admin, hospital admin, discharge communication) |
|  | Were you happy with how that communication went? |
|  | Did the people you communicated with listen to you and understand your needs? |
|  | Did they respond in the way you wanted? |
|  | Was there anything you particularly liked or disliked about the communication? |
| Individualised Care | How confident were you in your surgeon and their team? |
|  | Why did you feel confident or not confident? |
|  | Was there anyone in particular who influenced your confidence? |
| Psychological Recovery | What worried you most about your surgery? |
|  | Did you receive any help to address your concerns? |
|  | Did you feel you needed emotional or psychological support? |
|  | Were you offered or did you receive emotional or psychological support? |
| Coordination of Care | Did you feel your care was well-coordinated between different healthcare providers? |
|  | Were there any points where you felt that coordination between the different members of your care team could have been improved? |
| Return to Function | Did you have a clear goal of what you wanted to achieve after surgery (e.g., functional recovery)? |
|  | Was your recovery goal discussed with your healthcare team? |
|  | What assistance were you given in order to achieve your recovery goal? |
|  | Was your recovery followed up in a way that you found helpful? |
| Pain Management | Did you have a clear plan for managing your pain after surgery? |
|  | Were you informed of any potential side effects of the pain medication? |
|  | Were you given a plan to stop taking pain medication? |
|  | Were other forms of pain relief (besides medication) discussed with you? |
