## Supplementary File 2 for "Integrating patient experiences of healthcare: a critical realist thematic analysis of peri-operative experiences"

##### Supplementary File 2. ‘Big Q’ qualitative reporting guidelines (BQQRG) for authors, reviewers and editors.

| **Article section of element** | **Alignment with BQQRG guidelines** |
| --- | --- |
| Writing style | This study employs humanised and reflexive language to illustrate the emotional, psychological, and clinical dimensions of perioperative recovery. The writing prioritises patient narratives, using vivid, contextualised descriptions to foreground participants’ lived experiences and the interplay of key themes. Reflexivity is integrated throughout, positioning the researchers within the analytic process while maintaining a focus on the participants’ voices and experiences. |
| Terminology | This manuscript uses clear and consistent terminology, with key terms like 'psychological resilience' and 'care coordination' defined or operationalised in context. The language avoids pathologising, focusing on patient experiences and maintaining coherence throughout, aligning with the BQQRG guidelines. |
| Title | The title effectively captures the topic area and central storyline of the study. It includes key elements such as the methodology and thematic focus, while engaging the reader with its emphasis on psychological and emotional recovery. The inclusion of essential keywords ensures discoverability without relying on abbreviations |
| Abstract and keywords | The abstract conveys the study’s purpose, methodology, and key findings, highlighting its focus on emotional, psychological, and clinical dimensions of perioperative care. It is structured to engage readers while providing sufficient detail on themes, methods, and implications. The proposed keywords are directly aligned with the study’s central topics and enhance discoverability by including thematic, methodological, and conceptual terms. |
| **Introduction** | |
| Background and rationale | The introduction frames the research as part of a broader conversation on perioperative patient experiences, situating the study within existing gaps in the literature. It provides a robust rationale by discussing critical factors such as communication, emotional support, and care coordination, highlighting their interplay and underexplored nature. Contextualising the study within prior research, particularly the scoping review by Mickan et al. (2024), underscores the relevance of further exploring these themes. |
| Research question, aims, and/or purpose | The study clearly articulates its aim to explore perioperative patient experiences, building on pre-established themes while investigating emergent insights. It addresses the central research question of how emotional, psychological, and clinical dimensions converge to shape patient experiences, aligning with the Critical Realist paradigm. The purpose of amplifying patient voices and uncovering underlying mechanisms is consistent throughout. |
| ‘Owning your perspectives’ – discussion of theory | The introduction acknowledges the Critical Realist framework as central to the study’s design and analysis. It demonstrates theoretical congruence by linking the paradigm’s focus on observable phenomena and underlying mechanisms to the investigation of perioperative experiences. This grounding in theory provides a coherent lens through which the study examines both pre-established and emergent factors. |
| ‘Owning your perspectives’ – discussion of reflexivity | While reflexivity is primarily addressed in the methodology, the introduction implicitly acknowledges the researchers’ role in shaping the study focus. The emphasis on amplifying patient voices and recognising the emotional dimensions of perioperative care reflects a commitment to understanding how researcher perspectives influence the framing and interpretation of findings |

| **Methodology** |  |
| --- | --- |
| Research design/approach/methodology | The study employs a Critical Realist framework, utilising Christodoulou’s (2023) Four C’s model of thematic analysis to explore both observable perioperative experiences and underlying causal mechanisms. Semi-structured interviews provide a flexible yet structured approach to data collection, rooted in pre-established themes while allowing for emergent insights. The methodology demonstrates congruence between the study's aims, theoretical framing, and analysis, centring patient voices and embracing the complexity of their experiences |
| Participants/data sources/empirical materials | The study outlines participant selection criteria and recruitment processes, ensuring transparency and alignment with the study aims. Emphasis is placed on achieving dataset sufficiency through information power, prioritising the depth and richness of participant narratives over numerical size. Reflexivity is embedded, with the lead researcher acknowledging their clinical background and its potential influence. The flexible, participant-led interview approach fosters ownership and generates emotionally and contextually grounded data |
| Ethical approval and ethical code/principles followed | Ethical approval was obtained from the Bond University and all participants provided informed consent prior to participation. The study prioritised participant confidentiality through the use of pseudonyms and secure data storage. Reflexivity was embedded in the researcher-participant dynamics, with NM acknowledging the potential influence of his clinical background on interviews and analysis. Accessibility considerations were addressed by offering Zoom interviews, enabling participation regardless of location, and minimising barriers for eligible participants. |
| Data generation | Data generation employed semi-structured interviews, offering participants the flexibility to share narratives aligned with pre-established themes while allowing space for emergent insights. The conversational approach prioritised participant-led storytelling, fostering contextual richness and authentic expression. Reflexivity was embedded throughout, with the lead researcher critically reflecting on the potential influence of their clinical background. Data integrity was ensured through intelligent verbatim transcription and meticulous review of transcripts by multiple researchers. |
| Data analysis procedures | Thematic analysis followed Christodoulou’s Four C’s model, encompassing the systematic stages of coding, clustering, connecting, and constructing categories. This approach integrated pre-established themes and emergent factors, aligning with the study's Critical Realist paradigm. Retroductive reasoning enabled the exploration of causal mechanisms underlying perioperative experiences, while social network analysis visualised relationships between categories, enhancing depth, coherence, and interpretive rigour. |
| Quality practices | Rigour was upheld through reflexivity and peer debriefing, ensuring diverse perspectives informed the analysis. Thick descriptions captured the contextual richness of perioperative experiences, while Braun and Clarke’s conceptualisation of meaning generation as an evolving process informed the study's approach to data sufficiency. Methodological transparency and iterative collaboration reinforced the robustness of the findings. |
| **Analysis** | |
| Structure, number, and names of patterns / themes / categories / discourses / narratives (etc.) | The analysis balances pre-established and emergent categories, clearly delineating their structure and relational connections through a unified framework. Categories are presented with participant-centred names and contextual depth, highlighting their interplay within causal mechanisms. The approach avoids over-fragmentation, instead integrating categories into cohesive, multidimensional themes that reflect the complexity of perioperative experiences. |
| Overview of, or introduction to, analysis, if relevant | The analysis provides an overview of themes and categories, aligning closely with the analytic structure previewed in the methodology. Consistency in subheadings and theme names enhances clarity, while the addition of a thematic table contextualises the relationships between categories and patterns. |

| Theme/category/discourse/ narrative (etc.) conceptualisation, where relevant | The analysis integrates pre-established categories with emergent themes, maintaining coherence with the Critical Realist paradigm and Four C’s methodology. This approach ensures categories reflect both participant narratives and the broader study framework. Divergences between pre-established categories and emergent findings were critically evaluated and transparently addressed, enhancing alignment with the study's methodological rigor and patient experience focus. |
| --- | --- |
| Analytic depth | The study provides a detailed interpretive analysis, grounded in the Critical Realist paradigm, that contextualises data extracts within the perioperative experience. The emphasis on meaning-making avoids foreclosure, ensuring that emergent themes are presented as complementary to pre-established insights rather than reductive. This depth allows the interplay of emotional, clinical, and systemic dimensions to be thoroughly explored. |
| Use of data extracts, where relevant | Rich and contextualised participant quotes highlight the findings, carefully selected to represent the complexity of perioperative experiences. These quotes balance narrative depth with participant confidentiality and align with the Critical Realist framework by linking individual accounts to broader thematic interpretations. |
| Integration of analysis with existing research and theory | The analysis synthesises patient-reported experiences with existing frameworks (e.g., Mickan et al., 2024), creating a coherent intersection between theory and emergent insights. This integration reflects the Critical Realist commitment to connecting surface-level observations with underlying mechanisms, ensuring that the findings align with and expand upon established literature. |
| **Discussion/Conclusion** | |
| Analytic conclusions and contributions | The discussion synthesises the study's themes and causal mechanisms, highlighting their contributions to understanding patient experiences in perioperative care. The analysis connects these findings to existing theoretical and clinical frameworks, illustrating how the research advances knowledge in both scholarly and practical domains. The broader implications for improving patient-centred care are emphasised. |
| Study evaluation/reflection | This section critically reflects on the study's design, methodologies, and outcomes, acknowledging its contextual scope and potential limitations. Rather than presenting limitations as deficits, the reflection explores how the study’s context and methodologies shaped the findings. It also considers the transferability of insights to similar healthcare contexts and practices. |
| Researcher reflexivity | The role of the researcher in shaping the research process is explicitly acknowledged, with reflection on how their clinical and academic perspectives influenced data generation and interpretation. Reflexivity is integrated throughout, highlighting the co-constructed nature of the findings and reinforcing the alignment with the Critical Realist approach. |
| Implications for future research, policy and practice, where relevant | The study outlines actionable insights for clinicians and healthcare systems, prioritising holistic and personalised care approaches. It suggests targeted areas for future research to further explore underrepresented patient groups or contextual variations, ensuring evidence-based recommendations that extend the study's practical impact without overstating its generalisability. |
