## Supplementary File 3 for "Integrating patient experiences of healthcare: a critical realist thematic analysis of peri-operative experiences"

##### Supplementary File 3. Qualtrics questionnaire

| **Item** | **Question** |
| --- | --- |
| 1. | What is your age? |
| 2. | What is your postcode? |
| 3. | What is your gender? |
| 4. | Have you undergone a surgical procedure in the past 2yrs? |
| 5. | How do you generally describe your recent experience of surgery to your friends and family? |
| 6. | What type of hospital did your operation take place in? |
| 7. | How do you feel about the length of time you had to wait to have your surgery? |
| 8. | Do you feel that your expectations of surgery were met? |
| 9. | Throughout the surgery process do you feel the information you received was accurate? |
| 10. | Did you feel that communication with your healthcare team was consistent throughout the whole surgery process? |
| 11. | Do you feel that the care you received met your individual needs? |
| 12. | Was your pain adequately managed after your surgery? |
| 13. | Do you feel you received the required level of psychological support for your recovery after surgery? |
| 14. | Was the care you received from your treating team well-coordinated, from GP referral through to post-operative rehabilitation? |
| 15. | Were you able to return to your expected level of functioning after your surgery? |
| 16. | Is there anything else you would like to add? |
