## Supplementary File 4 for "Integrating patient experiences of healthcare: a critical realist thematic analysis of peri-operative experiences"

##### Supplementary File 4. Clustering of codes into predetermined categories

|  |  | **Codes** |
| --- | --- | --- |
| Categories | C1 – Realistic Expectations | ANC1, ANC10, ANC2, ANTC1, ANTC16, ANTC17, ANTC18, ANTC36, BEC1, BEC2, ERC1, GOC1, GOC10, GOC3, GOC9, GRC1, GRC2, GRC9, JAC1, JAC17, JEC1, JEC2, JEC20, JEC21, JEC3, JOC10, JOC3, JUC16, JUC17, LIC6, LOC1, MAC1, PEC1, PEC20, PEC5, SUC1, SYC1, SYC3, SYC31, SYC32, SYC44, SYC9, TEC1, TEC15, TEC7 |
|  | C2 – Accurate Information | BEC20, ANTC14, ANTC15, ANTC28, ANTC38, ANTC41, ANTC44, ERC2, ERC9, GOC2, GOC5, GRC11, GRC3, JAC3, JAC9, JEC15, JEC4, JEC5, JEC6, JOC12, JOC4, JUC10, JUC15, LIC10, LIC12, LIC13, LIC17, LIC2, LIC32, LIC4, LOC19, LOC2, LOC7, MAC11, MAC15, MAC2, MAC9, PEC12, PEC13, PEC16, PEC17, PEC2, PEC22, PEC27, PEC29, PEC33, PEC4, SIC12, SIC2, SUC16, SUC4, SYC16, SYC17, SYC2, SYC25, SYC43, SYC50, SYC51, SYC7, TEC10, TEC2, TEC6 |
|  | C3 – Consistent Communication | ANC16, ANC25, ANC28, ANC3, ANC4, BEC14, BEC15, BEC17, BEC7, BEC9, ERC10, ERC12, ERC13, ERC14, ERC17, ERC21, ERC3, GOC12, GOC4, GRC10, GRC6, JAC10, JAC12, JAC16, JAC6, JEC7, JOC11, JOC2, JOC8, JOC9, JUC11, JUC13, LIC14, LIC23, LIC27, LIC29, LIC36, LIC5, LOC17, LOC3, LOC6, LOC9, MAC13, MAC3, MAC32, MAC8, PEC19, PEC24, PEC28, PEC3, PEC7, PEC8, PEC9, SIC16, SIC3, SUC13, SUC17, SUC22, SUC23, SUC29, SUC7, SYC13, SYC15, SYC24, SYC45, SYC49, SYC53, SYC54, SYC6, SYC64, TEC12, TEC3, TEC5, TEC8 |
|  | C4 – Individualised Care | ANC11, ANC14, ANC7, ANC8, ANTC19, ANTC24, ANTC27, ANTC31, ANTC37, ANTC6, ANTC7, ANTC9, BEC18, BEC5, BEC8, ERC19, ERC7, GOC13, GOC16, GOC8, JAC11, JAC15, JAC19, JAC4, JEC9, JUC14, JUC2, JUC8, LIC18, LIC22, LIC25, LIC28, LIC9, LOC10, LOC16, LOC29, LOC30, LOC5, LOC8, MAC10, MAC14, MAC16, MAC17, MAC29, PEC15, PEC23, PEC25, PEC36, SIC15, SIC4, SIC9, SUC19, SUC2, SUC26, SUC30, SYC14, TEC17, TEC4 |
|  | C5 – Coordination of Care | ANC12, ANC13, ANC17, ANC5, ANC6, ANTC10, ANTC21, ANTC25, ANTC26, ANTC30, ANTC40, ANTC42, ANTC43, ANTC5, BEC6, ERC18, ERC22, ERC5, GOC14, GOC6, GRC4, GRC8, JAC20, JAC5, JEC16, JEC8, JOC5, JUC3, LIC24, LIC38, LOC12, LOC18, MAC6, MAC7, PEC11, PEC30, SIC10, SIC6, SUC14, SUC21, SUC5, SUC8, SYC10, SYC11, SYC12, SYC28, SYC33, SYC36, SYC42, SYC52, SYC8, TEC11, TEC13, TEC16, TEC18, TEC19, TEC22 |

|  | C6 – Psychological Recovery | ANC15, ANC23, ANC9, ANTC11, ANTC12, ANTC13, ANTC2, ANTC20, ANTC22, ANTC23, ANTC3, ANTC32, ANTC39, ANTC4, BEC12, BEC13, BEC19, ERC20, ERC6, ERC8, GOC11, GOC15, GRC7, JAC18, JAC7, JAC8, JEC10, JEC17, JEC19, JEC23, JOC7, JUC1, JUC12, JUC4, JUC9, LIC1, LIC20, LIC21, LIC26, LIC34, LIC35, LIC37, LOC13, LOC14, LOC20, LOC4, MAC18, MAC19, MAC23, MAC25, MAC26, MAC28, MAC33, MAC35, PEC10, PEC14, PEC18, PEC31, PEC32, PEC34, PEC6, SIC11, SIC5, SUC15, SUC24, SUC28, SUC3, SUC6, SYC18, SYC19, SYC20, SYC21, SYC23, SYC27, SYC34, SYC37, SYC38, SYC39, SYC4, SYC40, SYC41, SYC46, SYC47, SYC48, SYC55, SYC56, SYC57, SYC58, SYC59, SYC61, SYC62, SYC65, SYC66, TEC21, TEC23, TEC9 |
| --- | --- | --- |
|  | C7 – Return to Function | ANC22, ANTC29, ANTC8, BEC16, ERC11, GOC7, GRC5, JAC14, JAC2, JAC21, JEC11, JEC22, JOC1, JOC6, JUC6, JUC7, LIC11, LIC19, LIC3, LIC30, LIC33, LIC8, LOC15, LOC23, LOC24, LOC26, LOC27, LOC28, MAC21, MAC27, MAC30, MAC34, PEC21, PEC26, PEC35, SIC13, SIC7, SUC11, SUC12, SUC20, SUC25, SUC27, SYC22, SYC35, SYC60, SYC63 |
|  | C7 – Careful Monitoring Around Pain Management | ANC18, ANC19, ANC20, ANC21, ANC24, ANC26, ANC27, ANTC33, ANTC34, ANTC35, BEC10, BEC11, BEC3, BEC4, ERC15, ERC16, ERC4, JAC13, JAC22, JEC12, JEC13, JEC14, JEC18, LIC15, LIC16, LIC31, LIC7, LOC11, LOC21, LOC22, LOC25, MAC12, MAC20, MAC22, MAC24, MAC31, MAC4, MAC5, SIC14, SIC8, SUC10, SUC18, SUC9, SYC26, SYC29, SYC30, SYC5, TEC14, TEC20 |
