## Supplementary File 5 for "Integrating patient experiences of healthcare: a critical realist thematic analysis of peri-operative experiences"

##### Supplementary File 5. Clustering of codes into emergent categories

|  |  | Codes |
| --- | --- | --- |
| Categories | C – Patient Empowerment Through Education | NAC1, NAC2, NAC3, NAC4, NAC5, NANC1, NANC2, NBEC1, NERC1, NERC2, NANTC1, NANTC2, NANTC3, NANTC4, NANTC5, NANTC6, NANTC7, NANTC8, NANTC9, NANTC10, NANTC11, NANTC12, NANTC13, NANTC14, NANTC15, NANTC16, NANTC17, NANTC18, NANTC19, NANTC20, NANTC21, NANTC22 |
|  | C2 – Healthcare System Navigation Challenged | NAC6, NANC3, NBEC2, NBEC3, NBEC4, NGOC1, NGOC2, NLOC1, NLOC2, NMAC1, NMAC3, NMAC4, NMAC5, NMAC6, NNAC1, NNAC2, NNAC3, NNAC4, NNAC5, NNAC6, NNAC7 |
|  | C3 – Postoperative Emotional Support Needs | NAC7, NAC8, NERC3, NGOC3, NPEC1, NPEC2, NSIC1, NSUC1, NSYC1, NTEC1, NSYC10, NSYC11, NSYC12, NSYC13, NSYC14, NSYC15, NSYC16, NSYC17, NSYC18, NSYC19, NSYC20, NSIC3, NSIC4, NSIC5, NSYC4, NSYC5, NSYC6, NSYC7, NSYC8, NSYC9 |
|  | C4 – Information Accessibility and Transparency | NANC4, NBEC5, NBEC6, NERC4, NGRC1, NJEC1, NJEC2, NLIC1, NLIC2, NMAC2, NSYC21, NSYC22, NSYC23, NSYC24, NSYC25, NSYC26, NTEC4, NTEC5, NTEC6 |
|  | C5 – Social and Familial Influence on Care Decisions | NANC5, NERC5, NERC6, NGRC2, NGOC4, NJAC1, NJEC3, NSUC2, NSYC2, NSYC3, NJEC4, NJEC5, NJEC6, NJEC7, NJAC2, NJAC3, NJAC4, NJAC5 |
|  | C6 – Post-Surgical Complication Management | NANC6, NANC7, NERC7, NERC8, NPEC3, NGOC5, NJUC1, NLIC3, NSUC3, NTEC2, NJOC1, NJOC2, NJOC3, NGRC4, NGRC5, NGRC6, NGRC7, NGRC8, NPEC4, NJUC3, NJUC4, NJUC5, NJUC6, NJUC7, NJUC8, NLIC5, NLIC6, NLOC3, NLOC4, NLOC5, NLOC6, NLOC7, NLOC8, NLOC9, NLOC10, NLOC11, NLOC12, NLOC13 |
|  | C7 – Financial barriers and resource accessibility | NANC8, NERC9, NERC10, NGRC3, NGOC6, NJUC2, NSIC2, NSUC4, NLIC4, NTEC3, NGOC7, NGOC8, NGOC9, NSUC5, NSUC6, NSUC7, NSUC8, NSUC9, NSUC10 |
