## Supplementary File 6 for "Integrating patient experiences of healthcare: a critical realist thematic analysis of peri-operative experiences"

##### Supplementary File 6. Representative patient quotes categorised by key category

### a) Realistic expectations

These codes encapsulate the emotional weight and variance in patients’ expectations regarding their perioperative experiences. The stark difference between anticipated and actual recovery, as well as the emotional responses, are significant in representing this category

| **Code** | **Text Segment** | **Code Description** |
| --- | --- | --- |
| ANTC18 | "I didn’t expect it to be this traumatic." | Demonstrates the significant discrepancy between the patient’s expectations and the harsh reality of the surgery experience. |
| GRC9 | "They definitely wouldn't commit to what sort of vision I would have, other than I should have some! That was about as good as it could get." | Highlights the cautious and uncertain expectations set by medical professionals regarding recovery outcomes. |
| ANC10 | "I knew it was going to be a better experience than my first, so I think that probably made me very relaxed." | Reflects how a patient's past positive experience influences their expectations, leading to a sense of calm and confidence. |

### b) Accurate information

These codes offer a strong reflection of the challenges patients face in receiving and understanding accurate, actionable information pre- and post-surgery. Each one highlights a critical aspect of the communication gaps or successes that shape the perioperative experience.

| **Code** | **Text Segment** | **Code Description** |
| --- | --- | --- |
| GOC5 | "When we first consulted with the surgeon in Sydney, they sent me a whole selection of books, with the side effects, what to expect in the operation, what to expect after the operation." | Provision of comprehensive information resources by the surgeon’s office to Gordon, enhancing his understanding of the procedure and recovery. |
| ERC9 | "Yes. He said that it would hopefully calm down within a few weeks. Make an appointment for 8 weeks - if it hasn't calmed down in 8 weeks, come back and see us. But the information he gave...was wrong." | Surgeon's prognostic information and follow-up advice, which the patient found to be incorrect. |
| ANTC38 | "I couldn’t really remember the times for nil by mouth, which led to confusion." | Reflects the impact of her ADHD on processing pre-surgery instructions. |

### d) Consistent communication

These codes highlight the critical impact of consistent communication on the perioperative experience. They reflect how communication breakdowns can lead to frustration and erode trust, emphasising the importance of clear, timely, and transparent messaging from healthcare providers.

| **Code** | **Text Segment** | **Code Description** |
| --- | --- | --- |
| BEC7 | "They did look at possibly using a local anaesthetic, but then decided against that." | Bernard describes the surgical team’s consideration of different anaesthetic options, reflecting communication between the patient and healthcare team regarding the best course of action. |
| JAC6 | "He drew a diagram for me of all my bits and pieces and how they fit it in and what they were doing." | Detailed explanation by the surgeon using visual aids enhanced understanding of the surgical procedure. |
| SYC49 | "I was really happy with the dialogue I had with the surgeons prior and post to be honest." | Positive pre- and post-surgery communication with the surgeons. |

### e) Individualised care

These codes highlight the impact of individualised care in enhancing the perioperative experience. Personal allowances, time considerations, and a tailored approach help patients feel valued and ensure their specific needs are addressed with care and attention.

| **Code** | **Text Segment** | **Code Description** |
| --- | --- | --- |
| BEC18 | "I think the most important thing is to treat people as a person, not a number, and to be aware of everyone’s cultural and ethnic differences and backgrounds..." | Bernard emphasises the importance of personalised care that considers cultural and ethnic differences, stressing that patients should be treated as individuals. |
| GOC16 | "Before the surgery, the doctor told me I had to get these 2 bags of fluid. It was something that rehydrated you […] and sort of aids in recovery. […] I was the only one out of the whole group who’d had that." | Gordon received specific preoperative treatment to aid in recovery, highlighting differences in treatment protocols among patients. |
| SUC26 | "I did ring the surgeon up about that and he said, look, it will correct over time and, and it did." | Susan appreciated her surgeon’s reassurance that her limp would resolve over time. |

### g) Coordination of care

These codes emphasise the critical role that coordination of care plays in patient outcomes. From rushed discharges to a lack of post-operative support, these examples reflect the challenges of inadequate coordination, while also highlighting the positive impact of effective specialist referrals.

| **Code** | **Text Segment** | **Code Description** |
| --- | --- | --- |
| ANTC5 | "They said ‘time to go’... I was bleeding and still screaming." | Describes a rushed discharge experience despite the patient's distress, showing a lack of coordinated care and sensitivity. |
| JAC5 | "9 providers, but no one could give me any house cleaning." | Highlights the difficulties in securing post-operative home care, underscoring gaps in coordinated care during recovery at home. |
| GRC8 | "Someone at the [hospital name] put me onto a really good surgeon who specialises in cornea injuries and who did my cataract operation, so that was a real turning point." | Being referred to a specialist greatly improved the patient's care trajectory, showing effective coordination and the value of specialist referrals. |

### h) Psychological recovery

These codes capture the intense psychological challenges patients face after surgery, ranging from trauma and confusion to feelings of emotional overwhelm and severe mental health struggles linked to post-surgical complications.

| **Code** | **Text Segment** | **Code Description** |
| --- | --- | --- |
| ANTC3 | "I woke up screaming... I don’t know if I was in pain or just traumatised." | Describes her confusion between pain and trauma post-surgery, highlighting deep emotional distress following the operation. |
| GOC11 | "There was one chap who came the other week talking about wanting to commit suicide because he couldn’t get an erection to have sex." | Reveals the severe psychological impact that post-surgery complications can have on patients, highlighting the need for psychological support. |
| SYC21 | "I was a powder keg ready to explode." | Emotional breakdown, revealing unaddressed psychological stress, described through a vivid metaphor. |

### j) Return to function

These codes highlight the emotional weight of regaining independence and facing the logistical and financial challenges in accessing necessary care. The selected codes depict both the physical and financial hurdles patients face in returning to normal function.

| **Code** | **Text Segment** | **Code Description** |
| --- | --- | --- |
| LIC33 | "I suppose getting the function back. [...] being able to be independent again and being able to do things without having to ask my husband." | Linda identifies her primary concern post-surgery as regaining function and independence, particularly in performing everyday tasks. |
| PEC26 | "The first 6 weeks I needed a lot of help [...] and it was probably 3-6 months before I was really able to return to full capacity." | Penny discusses her lengthy recovery period and the support she needed, emphasising the time required to return to full function. |
| SIC13 | "Because then you're left to get the physio yourself, so you can't afford it, you can't go all the time." | Financial and logistical barriers to accessing necessary post-operative physiotherapy. |

### k) Careful monitoring around pain management

These codes depict the emotional highs and lows of pain management, from excellent care and attentive pain relief to delays and miscommunication. They highlight the critical role of careful monitoring in managing patient pain effectively.

| **Code** | **Text Segment** | **Code Description** |
| --- | --- | --- |
| ANC24 | "The nurses were on top of everything, my wound got checked several times, and... my wound was checked again [at the follow-up visit]." | Annabelle appreciated the thorough follow-up and monitoring of her surgical wound, which contributed to her positive recovery. |
| LIC15 | "He got the anaesthetist to do that [...] the nerve block doesn't wear off, so you don't feel pain for a longer period of time." | Linda explains the pain management technique used during surgery, which helped prolong pain relief. |
| SUC9 | "I was given so many drugs and I was vulnerable... but I would have liked to have had more choice about that." | Susan expresses frustration with the over-prescription of medications without considering her preferences or informing her about side effects. |

### m) Patient empowerment through education

These codes reflect how access to information and clear communication empower patients to make informed decisions about their care. They capture patients' emotional responses to feeling more in control and confident throughout their perioperative journey.

| **Code** | **Text Segment** | **Code Description** |
| --- | --- | --- |
| NANTC13 | "The doctor gave me a booklet, and I could read up about the operation and prepare myself for what to expect." | The patient describes how the educational booklet provided by her doctor helped her understand and prepare for the surgery, giving her confidence in the process. |
| NANTC1 | "I’ve got bulimia and I rotted all the teeth from my mouth." | The patients ongoing struggle with bulimia, which significantly impacted her health and contributed to the need for surgery. |
| NANTC7 | "The anaesthetist was nice and tried to calm me, but I was beyond reach." | Despite efforts by the anaesthetist to provide reassurance, Anna experienced significant emotional challenges that remained unresolved, underscoring the importance of tailored psychological support. |

### n) Healthcare system navigation challenged

These codes capture the frustrations patients face when navigating a complex and disjointed healthcare system. From long wait times to repeated transfers between departments, they underscore the emotional and logistical toll that systemic inefficiencies can have on patients.

| **Code** | **Text Segment** | **Code Description** |
| --- | --- | --- |
| NBEC2 | "Because of my background health and also from my work, I approach things and view things a lot differently from an ordinary patient. I'm a bit different." | Bernard describes how his unique background as a counsellor influenced his experience and perceptions, highlighting the variability of patient perspectives based on personal history. |
| NMAC3 | "The nurses were extremely good at what they did, but they couldn’t answer a lot of my questions." | Competent in clinical duties, the nursing staff lacked the ability to provide detailed information. |
| NNAC6 | "My short-term memory [has been affected]. I never used to do it before, but I've got that thing where you walk into a room and forget why you’re there." | Discusses cognitive changes post-operation, likely due to the trauma or extended anaesthesia. |

### p) Postoperative emotional support needs

These three codes reflect both immediate emotional needs and the potential for long-term psychological effects due to insufficient support during recovery. They emphasise the importance of emotional care for mitigating long-term impacts.

| **Code** | **Text Segment** | **Code Description** |
| --- | --- | --- |
| NSYC22 | "I am 100%... that’s me to a tee. It came to a head... I just collapsed. I howled uncontrollably." | Emotional breakdown revealing intense internal struggle and unmet psychological needs. |
| NSYC15 | "I think to go through this, you can’t do it alone." | Reflects the necessity of having a strong support network during recovery, underscoring the emotional burden of facing surgery without adequate support. |
| NERC3 | "No, and I wouldn't have done it anyway, because I don't want to be a sook." | Highlights how societal norms around pain tolerance prevented the patient from seeking necessary emotional and psychological support, leading to potential long-term impact. |

### q) Information accessibility and transparency

These segments illustrate the emotional impact of unclear, standardised communication and the challenges faced in advocating for personal needs and understanding healthcare costs. Improved accessibility to tailored information is essential for enhancing patient outcomes.

| **Code** | **Text Segment** | **Code Description** |
| --- | --- | --- |
| NERC4 | "I know that as nurses, we say the same spiel to everybody. It's only when there's something drastically different to normal that you’d say something different." | This highlights how standardised communication practices may fail to meet individual patient needs, pointing to the need for more personalised and transparent information. |
| NANC4 | "We didn’t feel like we had a lot of room to advocate for ourselves [in Malaysia]." | Reflects the patient’s past experience of feeling disempowered due to poor information accessibility, which contrasts with their current healthcare experience. |
| NJEC1 | "I guess you're almost certainly aware that the Medicare rebates available for physiotherapy and allied health in general are abysmal, like terrible." | Highlights the frustration caused by limited financial support for necessary rehabilitation services, emphasising the need for transparent communication about costs and rebates. |

### s) Social and familial influence on care decisions

These segments highlight the complex role of family and societal judgments in shaping healthcare decisions. They explore the emotional challenges patients face, including discomfort discussing health conditions, external judgments, and the influence of medical diagnostics on family-involved decision-making.

| **Code** | **Text Segment** | **Code Description** |
| --- | --- | --- |
| NJAC2 | "I didn’t want to talk about bowels. I hate bowels." | Demonstrates how personal discomfort in discussing health conditions influences social interactions and decision-making. |
| NJEC4 | "I was scared that I'd get judgment from that, like being like in public, and I did receive judgment for that..." | Reflects the emotional impact of societal and familial judgments on healthcare decisions, particularly around mobility and recovery. |
| NGRC2 | "But at that stage the PET scan showed it hadn’t gotten out […] and it was still contained." | Describes how diagnostic clarity influenced care decisions, with family members heavily involved in understanding the medical situation. |

### t) Post-surgical complication management

These segments demonstrate the emotional toll and logistical challenges patients experience when dealing with post-surgical complications, emphasising systemic inefficiencies, the crucial role of family, and frustration with poor communication from healthcare professionals.

| **Code** | **Text Segment** | **Code Description** |
| --- | --- | --- |
| NERC7 | "Eventually after working 9 and a half hours, they gave me some Endone." | Describes the lengthy wait time before receiving adequate pain management, highlighting hospital care inefficiencies. |
| NPEC3 | "My mum moved in with us for 6 weeks [...] and my husband took quite a bit of time off work." | Emphasises the significant role family plays in managing complications post-surgery, illustrating the emotional and logistical challenges faced by patients. |
| NJUC6 | "And I'd like to know why. But I wouldn't get an answer." | Reflects frustration with the lack of clear communication from healthcare professionals, which may exacerbate complications and anxiety post-surgery. |

### v) Financial barriers and resource accessibility

These segments highlight the impact of financial barriers and resource accessibility, where patient confidence, introversion, and resource constraints can hinder the pursuit of adequate care and support.

| **Code** | **Text Segment** | **Code Description** |
| --- | --- | --- |
| NERC9 | "No, I had no question because I thought I knew everything. I was nursing for 30 years." | Reflects a patient’s sense of confidence from prior knowledge, which may mask unmet financial and resource needs. |
| NJUC2 | "I'm terribly introverted, and I don’t like causing trouble, I don't like being the centre of attention." | Illustrates how personal traits like introversion can prevent patients from seeking the resources and financial help they need. |
| NLIC4 | "My attitude changed a lot to things. [...] I just think, well, if something happens, all you can do is get out of it the best you can." | Describes how limited financial resources forced a patient to adopt a more pragmatic approach to their healthcare. |
