## Supplementary File 7 for "Integrating patient experiences of healthcare: a critical realist thematic analysis of peri-operative experiences"

##### Supplementary File 7: Unified categories and factors from perioperative patient experiences

| **Pre-established Categories** | **Emergent Categories** | **New Unified Categories** |
| --- | --- | --- |
| C1 - Realistic Expectations | C1 - Patient Empowerment through Education | C1 – Patient Empowerment & Realistic Expectations [merged] |
| C2 - Accurate Information | C4 - Information Accessibility and Transparency | C2 – Accurate & Accessible Information [merged] |
| C3 - Consistent Communication | No clear equivalent | C3 – Consistent Communication [retained] |
| C4 - Individualised Care | No clear equivalent | C4 – Individualised Care [retained] |
| C5 - Coordination of Care | C2 - Healthcare System Navigation Challenged | C5 – Care Coordination & System Navigation [merged] |
| C6 - Psychological Recovery | C3 - Postoperative Emotional Support Needs | C6 – Psychological Recovery & Emotional Support [merged] |
| C7 - Return to Function | C6 - Post-Surgical Complication Management | C7 – Functional Recovery & Complication Management [merged] |
| C8 - Careful Monitoring around Pain Management | No clear equivalent | Keep as New C8 – Pain Management & Monitoring [retained] |
| n/a | C5 - Social and Familial Influence on Care Decisions | C9 – Social and Familial Influence [new] |
| n/a | C7 - Financial Barriers and Resource Accessibility | C10 – Financial Barriers & Resource Accessibility [new] |
