## Supplementary File 8 for "Integrating patient experiences of healthcare: a critical realist thematic analysis of peri-operative experiences"

##### Supplementary File 8: One-mode network matrix illustrating connections among unified categories

|  | C1 | C2 | C3 | C4 | C5 | C6 | C7 | C8 | C9 | C10 |
| --- | --- | --- | --- | --- | --- | --- | --- | --- | --- | --- |
| C1 | 1 | 1 | 1 | 1 | 0 | 1 | 1 | 0 | 0 | 0 |
| C2 | 1 | 1 | 1 | 1 | 1 | 1 | 0 | 1 | 0 | 0 |
| C3 | 1 | 1 | 1 | 0 | 1 | 1 | 0 | 0 | 0 | 0 |
| C4 | 1 | 1 | 0 | 1 | 1 | 1 | 0 | 1 | 0 | 0 |
| C5 | 0 | 1 | 1 | 1 | 1 | 1 | 1 | 1 | 0 | 0 |
| C6 | 1 | 1 | 1 | 1 | 1 | 1 | 1 | 1 | 1 | 0 |
| C7 | 1 | 0 | 0 | 0 | 1 | 1 | 1 | 1 | 0 | 1 |
| C8 | 0 | 1 | 0 | 1 | 1 | 1 | 1 | 1 | 0 | 0 |
| C9 | 0 | 0 | 0 | 0 | 0 | 1 | 0 | 0 | 1 | 1 |
| C10 | 0 | 0 | 0 | 0 | 0 | 0 | 1 | 0 | 1 | 1 |
