## Supplementary File 9 for "Integrating patient experiences of healthcare: a critical realist thematic analysis of peri-operative experiences"

##### Additional file 9. Category relationships: detailed explanation

**C1 – Patient Empowerment & Realistic Expectations:**

- **C1 ↔ C2 (Accurate & Accessible Information)**: Patient empowerment relies on accurate and accessible information. Patients equipped with proper knowledge are more likely to have realistic expectations about their treatment, post-surgical outcomes, and recovery. Information clarity allows patients to actively engage in shared decision-making, contributing to their empowerment.
- **C1 ↔ C3 (Consistent Communication)**: Consistent communication ensures that the empowering information patients receive is reliable and timely. When healthcare providers communicate clearly, patients are empowered to manage their recovery with confidence and have realistic expectations about the process.
- **C1 ↔ C4 (Individualised Care)**: Patient empowerment fosters better individualised care. Educated and informed patients are more likely to express their preferences and needs, enabling healthcare providers to create a care plan tailored to their unique circumstances, aligning expectations with individual health conditions.
- **C1 ↔ C6 (Psychological Recovery & Emotional Support)**: Empowerment also directly impacts psychological recovery. When patients have realistic expectations and feel in control of their health journey, they tend to cope better emotionally, mitigating anxiety and fear associated with surgery and recovery.
- **C1 ↔ C7 (Functional Recovery & Complication Management)**: Empowered patients are more proactive in managing their functional recovery. By having realistic expectations, they are more prepared for the physical demands of rehabilitation and better equipped to manage potential complications.

**C2 – Accurate & Accessible Information:**

- **C2 ↔ C1 (Patient Empowerment & Realistic Expectations)**: Accurate information is foundational to patient empowerment, enabling patients to set realistic expectations for their care and recovery.
- **C2 ↔ C3 (Consistent Communication)**: For information to remain accurate and accessible, it must be communicated consistently across healthcare providers. Consistency ensures that patients receive coherent information at every stage of their care, reducing confusion and uncertainty.
- **C2 ↔ C4 (Individualised Care)**: Information accuracy enhances individualised care by ensuring that patients' treatment plans are based on reliable data specific to their needs and health conditions. Accurate information also allows providers to adapt care plans when necessary.
- **C2 ↔ C5 (Care Coordination & System Navigation)**: Healthcare system navigation is heavily dependent on the availability of accurate information. Patients must understand how to access care, navigate insurance and billing systems, and schedule follow-up appointments. Clear, accessible information simplifies this process.
- **C2 ↔ C6 (Psychological Recovery & Emotional Support)**: Accurate information reduces anxiety and enhances emotional recovery by giving patients the knowledge they need to understand their progress and potential challenges. Knowing what to expect helps patients manage emotional distress.
- **C2 ↔ C8 (Pain Management & Monitoring)**: Information plays a crucial role in effective pain management. Patients need to be informed about their pain management strategies, including medication use, side effects, and alternative therapies. Accurate information ensures that patients manage their pain appropriately, which is essential for recovery.

**C3 – Consistent Communication:**

- **C3 ↔ C1 (Patient Empowerment & Realistic Expectations)**: Communication consistency is critical for maintaining realistic expectations. When all healthcare providers convey the same messages, patients can rely on the information they receive, helping them stay empowered and informed throughout their care journey.
- **C3 ↔ C2 (Accurate & Accessible Information)**: Communication consistency ensures that accurate information is conveyed effectively. If communication is inconsistent or fragmented, the information patients receive may be contradictory, leading to confusion or misaligned expectations.
- **C3 ↔ C5 (Care Coordination & System Navigation)**: Effective communication between healthcare professionals is essential for care coordination. Consistent communication prevents misunderstandings and miscommunication, which can hinder the coordination of services and impact patient outcomes.
- **C3 ↔ C6 (Psychological Recovery & Emotional Support)**: Consistent communication is critical for providing emotional support to patients. Regular, clear communication from the healthcare team can help reassure patients, address their concerns, and improve their psychological resilience during recovery.

**C4 – Individualised Care:**

- **C4 ↔ C1 (Patient Empowerment & Realistic Expectations)**: Individualised care empowers patients by providing treatment that reflects their specific needs and circumstances. Patients with realistic expectations are better able to engage with their care team and participate in shared decision-making, leading to more personalised care.
- **C4 ↔ C2 (Accurate & Accessible Information)**: Personalised care relies on having accurate and accessible information about the patient’s health status, medical history, and preferences. Tailoring care plans requires a flow of reliable information between patients and healthcare providers to ensure that treatments are aligned with individual needs.
- **C4 ↔ C5 (Care Coordination & System Navigation)**: Coordinating individualised care involves multiple healthcare professionals, and effective communication and information-sharing are essential. Coordination ensures that all members of the care team are working together to meet the patient’s specific needs, navigating the complexity of healthcare systems effectively.
- **C4 ↔ C6 (Psychological Recovery & Emotional Support)**: Individualised care also extends to psychological support, as emotional well-being is often deeply personal. Tailored psychological interventions, such as counselling or therapy, contribute to the overall mental recovery of patients, ensuring that psychological recovery is integrated with the physical aspects of care.
- **C4 ↔ C8 (Pain Management & Monitoring)**: Personalised pain management is crucial in individualised care, ensuring that pain relief strategies are tailored to the patient’s specific condition and tolerance. This helps patients engage fully in rehabilitation without being hindered by unmanaged or poorly controlled pain.

**C5 – Care Coordination & System Navigation:**

- **C5 ↔ C2 (Accurate & Accessible Information)**: Coordination of care relies on having access to up-to-date and accurate information. Care professionals must share patient records, treatment plans, and other critical information to ensure that the patient receives seamless care from multiple providers.
- **C5 ↔ C3 (Consistent Communication)**: Consistent communication is key to successful care coordination. All members of the healthcare team must communicate regularly and clearly to ensure that the patient’s care is coherent and continuous.
- **C5 ↔ C4 (Individualised Care)**: Coordinated care is necessary to deliver individualised care effectively. Healthcare providers must work together to ensure that the care plan is personalised to the patient’s specific needs and that every professional involved is aligned with the overall treatment goals.
- **C5 ↔ C6 (Psychological Recovery & Emotional Support)**: Coordinated care that includes psychological support ensures that emotional recovery is integrated into the patient’s overall treatment plan. Psychological resilience is necessary for navigating complex healthcare systems and managing the emotional strain of recovery.
- **C5 ↔ C7 (Functional Recovery & Complication Management)**: Coordinating care for functional recovery and complication management ensures that patients receive the appropriate follow-up care, rehabilitation, and interventions necessary for a successful recovery.
- **C5 ↔ C8 (Pain Management & Monitoring)**: Pain management often requires input from multiple healthcare professionals, including surgeons, nurses, and anaesthetists. Coordinating pain management strategies helps prevent gaps in care and ensures that the patient’s pain is consistently monitored and treated.

**C6 – Psychological Recovery & Emotional Support:**

- **C6 ↔ C1 (Patient Empowerment & Realistic Expectations)**: Psychological recovery is facilitated when patients are empowered with realistic expectations about their treatment and recovery. Patients with a clear understanding of the challenges ahead are more resilient in handling emotional distress and psychological adjustments.
- **C6 ↔ C2 (Accurate & Accessible Information)**: Clear and accessible information reduces uncertainty and anxiety, playing a crucial role in psychological recovery. Well-informed patients are better able to mentally prepare for the physical demands of recovery, reducing emotional stress and promoting mental well-being.
- **C6 ↔ C3 (Consistent Communication)**: Consistent communication between healthcare providers and patients ensures that psychological support is maintained throughout the perioperative journey. Regular updates and a reliable flow of information help alleviate anxiety, contributing to emotional stability and recovery.
- **C6 ↔ C4 (Individualised Care)**: Personalised psychological interventions cater to the emotional and mental health needs of each patient. By addressing individual psychological recovery processes, healthcare providers can better support patients in managing their emotional well-being alongside their physical recovery.
- **C6 ↔ C5 (Care Coordination & System Navigation)**: Coordinated care ensures that emotional and psychological needs are addressed across different providers, ensuring continuity of mental health support. This holistic approach facilitates smoother psychological recovery by providing a seamless integration of physical and mental healthcare services.
- **C6 ↔ C7 (Functional Recovery & Complication Management)**: Psychological resilience is key to managing post-surgical complications and progressing toward functional recovery. Patients with strong psychological support are more motivated to adhere to rehabilitation plans and overcome physical challenges, resulting in better functional outcomes.
- **C6 ↔ C8 (Pain Management & Monitoring)**: Pain management is closely linked to emotional well-being, as uncontrolled pain can lead to increased psychological distress. By ensuring pain is managed effectively, healthcare providers help stabilise emotional health, promoting smoother psychological recovery.
- **C6 ↔ C9 (Social & Familial Influence on Care Decisions)**: Social and familial support systems are essential for psychological recovery, providing emotional backing that helps patients cope with the stresses of recovery. Family involvement often alleviates psychological strain, creating a supportive environment for mental health recovery.

**C7 – Functional Recovery & Complication Management:**

- **C7 ↔ C1 (Patient Empowerment & Realistic Expectations)**: Empowered patients with realistic expectations are better prepared to manage post-surgical complications and achieve functional recovery. Understanding the recovery process empowers patients to take an active role in their rehabilitation, leading to more successful physical outcomes.
- **C7 ↔ C5 (Care Coordination & System Navigation)**: Coordinated care ensures that post-surgical complications are addressed promptly, supporting the patient’s functional recovery. By aligning the efforts of all healthcare providers involved, potential complications are managed more efficiently, enabling smoother rehabilitation.
- **C7 ↔ C6 (Psychological Recovery & Emotional Support)**: Psychological resilience is a critical factor in managing complications and achieving functional recovery. Patients who receive adequate emotional support are more likely to stay motivated and engage fully in their rehabilitation, leading to improved physical recovery.
- **C7 ↔ C8 (Pain Management & Monitoring)**: Effective pain management is vital for functional recovery, as poorly controlled pain can hinder a patient’s ability to participate in rehabilitation exercises. Ensuring that pain is managed properly allows patients to engage in physical activity, speeding up the recovery process.
- **C7 ↔ C10 (Financial Barriers & Resource Accessibility)**: Financial barriers can significantly impact a patient’s ability to access the resources necessary for functional recovery. Without adequate financial support, patients may struggle to afford rehabilitation services or essential post-surgical care, delaying their return to function.

**C8 – Pain Management & Monitoring:**

- **C8 ↔ C2 (Accurate & Accessible Information)**: Patients must receive clear information about their pain management options, medications, and potential side effects. Accurate information ensures that patients understand how to manage their pain effectively.
- **C8 ↔ C4 (Individualised Care)**: Pain management must be tailored to the patient’s specific needs. Individualised pain management plans help address each patient’s unique pain threshold, medical history, and recovery trajectory.
- **C8 ↔ C5 (Care Coordination & System Navigation)**: Pain management strategies must be coordinated across all healthcare providers to ensure consistent monitoring and care. Effective coordination ensures that pain management is continuously evaluated and adjusted as needed to meet the patient's needs.
- **C8 ↔ C6 (Psychological Recovery & Emotional Support)**: Pain management is directly linked to emotional well-being. Patients who experience uncontrolled pain are more likely to suffer from psychological distress, such as anxiety and depression. Conversely, well-managed pain can reduce emotional strain, improving psychological recovery.
- **C8 ↔ C7 (Functional Recovery & Complication Management)**: Effective pain control is essential for functional recovery, as patients are more likely to engage in rehabilitation activities when their pain is manageable. Uncontrolled pain can hinder physical progress and delay functional recovery.

**C9 – Social and Familial Influence on Care Decisions:**

- **C9 ↔ C6 (Psychological Recovery & Emotional Support)**: Social and familial support plays a significant role in emotional recovery. Patients who receive emotional and practical support from family members are better able to cope with the psychological challenges of post-surgical recovery. Family involvement can provide motivation and reassurance, helping patients navigate the mental and emotional aspects of recovery.
- **C9 ↔ C10 (Financial Barriers & Resource Accessibility)**: Family and social networks often help patients overcome financial challenges by providing emotional and financial support. Families may assist with navigating insurance issues, covering medical costs, or accessing necessary resources that patients may struggle to afford on their own.

**C10 – Financial Barriers & Resource Accessibility:**

- **C10 ↔ C7 (Functional Recovery & Complication Management)**: Financial barriers can directly impact a patient's ability to manage post-surgical complications and engage in functional recovery. Patients without access to sufficient financial resources may struggle to afford necessary follow-up care, medications, or rehabilitation services, which can hinder their recovery.
- **C10 ↔ C9 (Social and Familial Influence on Care Decisions)**: Family and social networks can alleviate some of the financial burden by offering emotional, logistical, and financial support. Family members may help patients navigate financial challenges, such as accessing insurance benefits or securing funding for necessary treatments
